# Consecutive day effects between sleep quality and affective symptoms among youth in the Brazilian High-Risk Cohort study

**DOI:** 10.64898/2026.07.14.26358099

**Authors:** Mathew R Varidel, Luke J Borgnolo, Victor An, Joanne S Carpenter, Ian B Hickie, Pedro M Pan, Francisco da Silva, Jacob J Crouse, Eurípedes C Miguel, Luís A Rohde, Giovanni A Salum, Frank Iorfino

## Abstract

**Background:** Bidirectional next-day associations between sleep disturbances and affective symptoms have been shown in previous research, yet the consecutive day effects between these factors remains poorly understood.

**Methods:** We analysed longitudinal ecological momentary assessment (EMA) data obtained from a subsample of young persons in the Brazilian High-Risk Cohort (BHRC) study collected in 2020-2021. Participants reported sleep quality each morning and rated affective symptoms relating to mood, anxiety, and energy four times daily for 28 days. We selected 88 individuals (17.83±1.74 years, 56 [63.6%] female gender) with at least one instance where individuals were observed three-days in a row. Within-person bidirectional next-day effects between sleep quality and affective symptoms were estimated using mixed-effects regression analysis adjusting. We then applied g-estimation approaches to estimate the effect that lagged sleep quality and consecutive improvements in sleep quality had on affective symptoms.

**Results:** Sleep quality and affective symptoms had bidirectional next-day effects, with sleep quality tending to have greater influence on affective symptoms than the reverse. Improved lagged sleep quality had positive effects on affective symptoms incrementally above the prior night’s sleep quality. Also, improvement of sleep quality across consecutive days had incremental and approximately equal effects on affective symptoms.

**Conclusions:** Sleep quality and affective symptoms exhibit a feedback loop, whereby poor sleep quality influences affective symptoms over consecutive days. Breaking these feedback loops, by improving sleep quality across several consecutive nights should improve affective symptoms. This supports interventions that target sustained improvement in sleep and possibly circadian regulation to improve affective symptoms.

## 1. Introduction

The relationship between sleep disturbances and negative affect is well established. Bidirectional associations have been reported in longitudinal studies (Alvaro et al., 2013; Difrancesco et al., 2021; Talbot et al., 2012; Van Dyk et al., 2016; Yasugaki et al., 2025) and evidence for interventions targeting sleep leading to significant positive effects on mental health in a dose-response relationship (Scott et al., 2021). Circadian rhythm disturbances have been identified as a potential mechanism underlying sleep disturbances in this context (Crouse et al., 2021), suggesting that sleep and circadian disturbances are a key intervention target in the treatment of mental ill-health (Freeman et al., 2020; Harvey et al., 2011; Meyer et al., 2024). Within this framework, consecutive nights of sleep disturbances could be driven by circadian dysregulation (i.e., aberrant strength and/or timing of sleep and wake signals), which could potentially influence mood disturbances.

When studying sleep disturbance, subjectively rated sleep quality is a practical measure that can be captured relatively easily by a single question (Snyder et al., 2018) and reflecting not only duration of sleep, but also latency, efficiency, and nighttime awakenings (Nelson et al., 2022). Poorer sleep quality has been associated with increased variability in sleep-wake patterns (Bei et al., 2016). Further, sleep quality tends to have greater next-day associations with affective symptoms than objective or subjective sleep duration (Difrancesco et al., 2021), which may be due to subjective sleep quality acting as a mediating factor in the relationship between variability in sleep duration and negative affect (Bei et al., 2017). Both poorer sleep quality and greater variability or irregularity in sleep are associated with misaligned or disrupted circadian rhythms (Hartstein et al., 2025; Mentzelou et al., 2023), suggesting that sleep quality may be a useful proxy for underlying sleep-wake and circadian disturbances.

Traditional research approaches relying on retrospective self-reports or laboratory sleep studies to examine associations between sleep quality and affect may fail to capture the nuanced temporal patterns that unfold in naturalistic ‘real-world’ settings. Alternatively, Ecological Momentary Assessment (EMA) methodologies, leveraging digital technologies to collect real-time data in participants’ natural environments, offer a powerful approach to examine these relationships with greater ecological validity (Shiffman et al., 2008). EMA protocols using smartphone-based assessments enable researchers to capture daily variations in affect and sleep while minimizing recall bias. This approach has demonstrated significant advantages over traditional retrospective methods, revealing that the relationship of sleep with mood and affect is more complex than previously understood. Previous EMA studies examining sleep-mood dynamics have identified bidirectional associations (Hickman et al., 2024) with significant immediate effects of sleep quality on next-day mood and affective symptoms (Blaxton et al., 2017; Bower et al., 2010), and evidence suggesting these effects may be stronger than the influence of mood and affective symptoms on subsequent sleep. However, whether these effects accumulate over consecutive days or dissipate following recuperative sleep has not yet been examined.

Research examining consecutive nights of sleep loss and mood show varied findings across studies, highlighting the complexity of day-to-day sleep-mood dynamics. Dinges et al. (1997) demonstrated cumulative negative effects on subjective sleepiness and mood (including fatigue, confusion, tension, and total mood) across seven nights with subjective changes often preceding objective performance decrements when sleep duration was experimentally restricted to approximately 5 hours per night. Importantly, they observed that these effects were not steady state but rather escalated over time, with pronounced changes appearing both early (days 1-2) and late (days 6-7) in the sleep restriction period. They also observed that complete recovery from these effects appeared to require two full nights of sleep, suggesting the accumulation of a “sleep debt” with corresponding mood consequences. By contrast Booth et al. (2021) reported that two nights of recovery of sleep duration within an experimental setting was insufficient for adolescents to recover from negative mood states following nine nights of 5-hour sleep restriction. Similarly, Lee (2022) showed that naturally occurring loss of sleep duration is associated with poorer consecutive-day affective symptom trajectories using observational diary data.

More recent work in healthy young males did not find effects of moderate sleep restriction (5-7 hours in bed) over seven consecutive nights on overall mood disturbance, with only poorer fatigue-inertia associated with the most restricted condition (Harous et al., 2021). Sleep deprivation can also have rapid but transient antidepressant effects (Giedke and Schwäzler, 2002), further complicating these day-to-day dynamics. These conflicting findings highlight a need to further explore the cumulative effects of sleep disturbance on affective symptoms.

Sleep quality also remains unexplored in this context. Critical gaps remain in understanding how consecutive nights of poor subjective sleep quality, as opposed to experimentally controlled sleep restriction, impact emotion regulation capabilities and mood states in naturalistic settings. Furthermore, the potential for individual differences in vulnerability to consecutive poor sleep—based on factors such as age, gender, or pre-existing mental health conditions—represents an important area for investigation.

The present study employs a smartphone-based EMA methodology to examine the temporal dynamics between consecutive nights of poor subjective sleep quality and daily mood fluctuations in a naturalistic setting among youth. By modelling the influence of consecutive poor sleep on mood, this research aims to provide a more nuanced understanding of sleep’s role on affective symptoms to inform targeted sleep interventions for improving mental health.

## 2. Methods

### 2.1 Data

Participants were recruited from the Brazilian High-Risk Cohort study (Salum et al., 2025, BHRC, 2015). BHRC is an ongoing multi-centre longitudinal study that follows 2511 children and adolescents born between 1998 and 2004 in Porto Alegre and São Paulo, Brazil. The BHRC study was approved by the Brazilian National Ethics in Research Commission (CAAE 13852413010015327 and 74563817.7.1001.5327). For screening, Wave 0, Wave 1, and Wave 2, written consent was obtained from parents and, when appropriate, assent or consent from participating children. For those unable to read or write, verbal agreement was documented after a researcher had explained the procedures and answered questions. Families could schedule appointments with study psychologists and social workers to receive results and referrals when indicated. By Wave 3, participants had reached the legal age of consent; consent was obtained directly from them. For participants lacking legal capacity, consent was obtained from parents/guardians with verbal assent from the participant. Situations involving risk of harm received special attention that was consistent with ethical guidelines.

A subsample of these individuals were recruited to complete a study using EMA from April 2020 to May 2021, which includes a period affected by COVID-19. They were asked to complete a morning questionnaire plus four EMAs each day for 28 days. In the morning, participants were asked about their sleep quality on a 1 to 5 Likert scale, where higher values correspond to better sleep quality (“How would you rate the quality of your sleep? Very poor, Poor, Fair, Good, Very good”). At each EMA individuals were asked several questions, mainly from the mMARCH EMA protocol (Dunton et al., 2014) from which we selected questions asking about momentary anxiety (relaxed vs. anxious: “Very relaxed, Relaxed, Somewhat relaxed, Neutral, Somewhat anxious, Anxious, Very anxious”), irritability (calm vs. angered: “Very calm, Calm, A little calm, Neutral, A little angry, Angry, Very angry”), sadness (happy vs sad: “Very happy, Happy, A little happy, Neutral, A little sad, Sad, Very sad”), overall mood (“How do you feel right now? 3 thumbs up, 2 thumbs up, 1 thumb up, neutral, 1 thumb down, 2 thumbs down, 3 thumbs down”), inactivity (active vs. inactive: “Very active, Active, A little active, Neutral, A little inactive, Inactive, Very inactive”), and tiredness (energetic vs. tired: “Very energetic, Energetic, A little energetic, Neutral, A little tired, Tired, Very tired”). We refer to these as ‘affective symptoms’ throughout this article. Each question relating to affective symptoms is asked on a 1 to 7 Likert scale, where higher values correspond to poorer symptoms. Example of the question interface is shown in Fig. S1.

We also use a set of ‘control variables’ for each analysis, with the intention of adjusting associations to have greater causal interpretation. First, within the BHRC data there is a high-risk grouping that we adjust for throughout this analysis. The high-risk grouping was based on high levels of symptoms within the family assessed by the family liability index (Salum et al., 2015). We also adjust for age, gender (female vs. male), study site (Porto Alegre vs. São Paulo) and weekend day.

### 2.2. Statistical Analysis

Our analysis targeted the within-person total effects between sleep quality and each of the six affective symptoms. Mathematical detail for our estimation procedure can be found in Supplementary Material 2; however, we will provide a high-level description here. We used causal inference approaches to strengthen the interventional interpretation of our estimates (Hernán and Robins, 2020; Pearl et al., 2016). The goal of our analysis is to provide estimates of the within-person effect of an exposure on an outcome in the case where the exposure was intervened upon, improving its relevance for clinical application (Prosperi et al., 2020; Sanchez et al., 2022). In practice, this was achieved by estimating mixed-effects linear regression models adjusting for prior day observations. Also, random intercepts for the outcome and individual-level mean centring for each exposure were introduced to allow for the estimation of within-person effects (Hamaker, 2025). Note within our approach, the time-invariant control variables (high-risk grouping, age, gender, study site) were incorporated to improve estimation of the random intercept.

First, we estimated next-day bidirectional effects between sleep quality and affective symptoms. When estimating the total effect of sleep quality on a next-day affective symptom, we adjusted for the prior day’s affective symptoms along with the control variables. When estimating the influence of an affective symptom on subsequent night’s sleep quality, we adjusted for all lagged affective symptoms, along with the control variables, broadly following a similar approach by Triantafillou et al. (2019). The reported average treatment effect (ATE) corresponds to the adjusted fixed-effect regression coefficient between the exposure and outcome.

Second, we examined whether lagged sleep quality had an additional effect on each affective symptom, beyond the prior night’s sleep quality. We used a two-step procedure following Loh and Ren (2023). First, we remove the portion of current-day affective symptom explained by the prior night’s sleep quality. Then we regressed that adjusted current-day affective symptom on lagged sleep quality, controlling for twice-lagged sleep quality and mood symptoms.

Third, we estimated the expected impact of different interventional sleep quality scenarios across two consecutive nights. We used a consecutive-day simulation-based approach to do this based on multivariate regression analyses, once again adjusting for prior day observations and the control variables. Our approach involves estimating the sleep-affect dynamics day-to-day, and then simulating an idealised intervention by adjusting sleep quality on consecutive nights (Pearl et al., 2016). We compared four scenarios. A ‘stable good sleep’ scenario, where sleep quality is set to the best possible value each day (on the 1-5 Likert scale). A ‘stable poor sleep’ scenario, where sleep quality is set to the lowest value on both days. We then investigate two mixed interventions. First, an ‘improving sleep’ scenario, where lagged sleep quality is set to the lowest value followed by the best. Second, a ‘worsening sleep’ scenario, where lagged sleep quality is set to the best followed by the lowest.

As individuals had varying numbers of observations, we estimated random slopes model for each estimation. Uncertainties given the varying numbers of observations were carried forward due to this hierarchical approach.

### 2.3. Statistical Estimation

All statistical analyses were performed in R (version 4.4.2, R Core Team, 2024). We estimate parameters within the Bayesian framework, with posterior sampling performed using high-level functionality to the underlying *stan* package (Bürkner, 2017; Goodrich et al., 2024). Stan is a probabilistic programming language that allows for posterior sampling using a Hamiltonian Monte Carlo procedure. We retained default weakly informative priors outlined in these packages documentation. We estimated each model using four chains with 2000 posterior samples after warmup. In accordance with previous recommendations, we ensured that the chains converged to similar estimates (i.e., 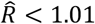) and had an appropriate number of effective sample sizes (at least 100 effective samples per chain) such that standard quantities (e.g., mean, standard deviation, etc.) could be estimated with reasonable accuracy (Vehtari et al., 2021).

Our procedures require several estimation steps. We ensure that uncertainties are passed through at each step by simulating uncertain quantities. These simulated quantities are then used to estimate the next step.

For all estimands, we report the mean and standard error within the main body of the article. Where possible, we discuss levels of evidence in a continuous way but highlight two thresholds, corresponding to a two-sigma (i.e., posterior probability, p < 0.05) and three-sigma (i.e., p < 0.003) level of evidence. To aid this discussion, we note that a strict Bonferroni correction assuming 24 independent tests, and a family-wise alpha of 0.05, would correspond to a corrected alpha of approximately 0.002, roughly corresponding to a three-sigma threshold. We get to 24 tests due to 6 each for analysis: 1) sleep quality on affective symptoms, 2) affective symptoms on sleep quality, 3) lagged sleep quality on affective symptoms, and 4) conseceutive nights sleep quality on affective symptoms.

### 2.4. Selection Criteria

We used a hybrid completed-cases approach to data selection. Self-reported daily affective symptoms were estimated by calculating the mean of observed symptoms across a day. No imputation was performed for sleep quality. All completed days were then retained. There are three analyses outlined above which require differing numbers of consecutive days to be completed. For the next-day effects, we required two consecutive days. Whereas, for the consecutive day analyses we required three consecutive days. For the sequential scenarios, we also remove rows with overlapping days to avoid double counting of data.

## 3. Results

### 3.1. Selection criteria and sample

We retained the maximum number of individuals for each analysis. The selection procedure is graphically outlined in Supplementary Material 3. For the final consecutive day analysis, we selected 88 individuals with mean age 17.83 years (SD, 1.74 years, min., 13.85, max., 22.48) of which 56 (63.6%) were female gender (Table 1). We show their baseline characteristics measured at the first day of EMA observations in Table 1. This is compared to those that had a baseline observation but were not selected in the analysis (n=183) where 100 (54.6%) were female gender. We checked for differences in means and proportions of demographics and EMA variables across the included and excluded groups. We did not perform any correction to account for our inclusion criteria as the difference for each variable were negligible. All estimated differences lie within the two standard deviations of zero and corresponding p-values ≥ 0.10. For completeness, for the day-to-day sleep-affect analysis, we selected 134 individuals with a median of three observations per person (min., 1, Q1, 1, Q3, 6, max., 28). For the influence of lagged sleep quality on affect, we had 88 individuals with 454 observations.

**Table 1.**
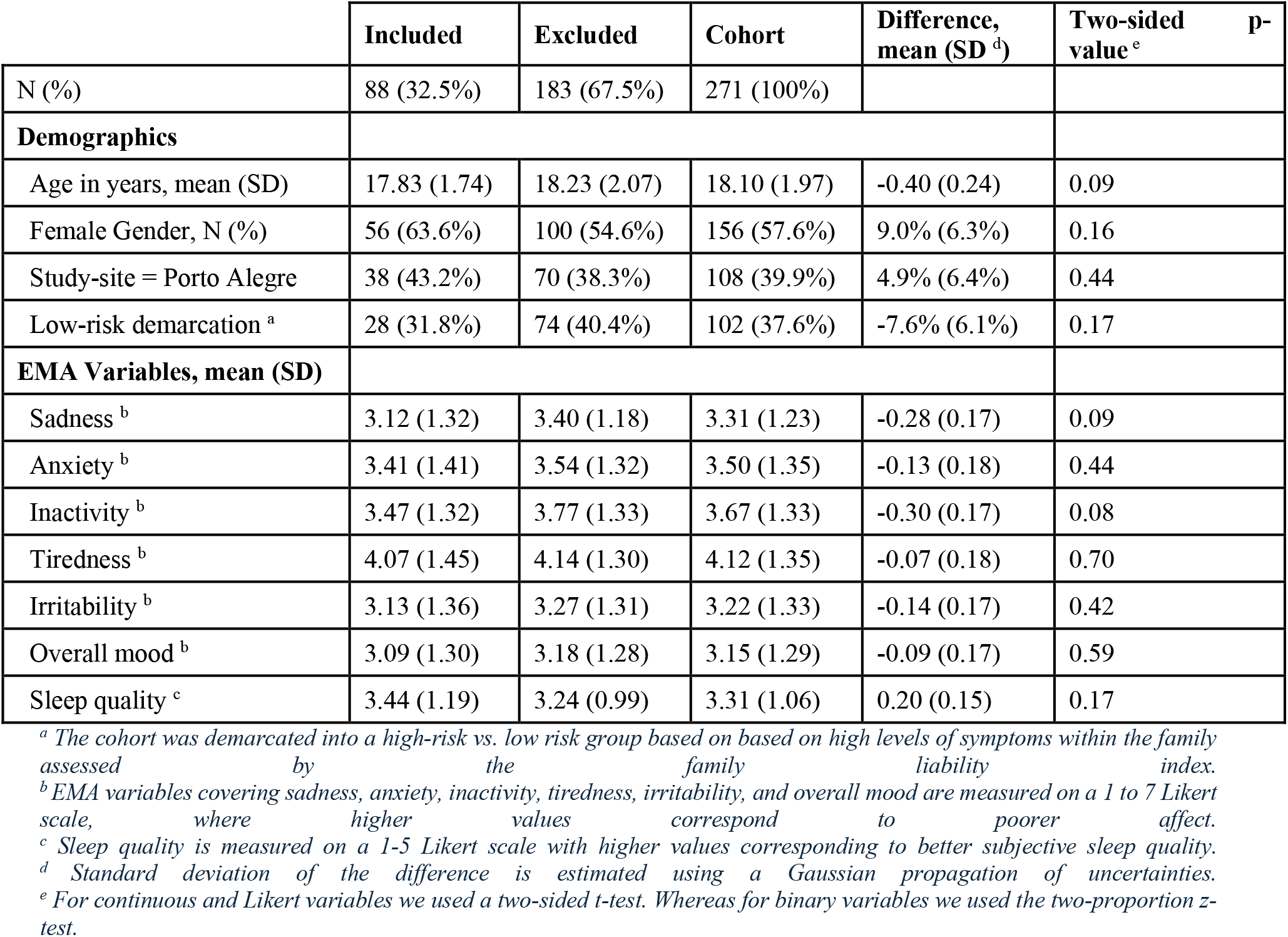
Sample characteristics for demographics and first-day averaged observations for those included within the analysis compared to those excluded.

|  | Included | Excluded | Cohort | Difference, mean (SD <sup>d</sup>) | Two-sided p-value <sup>e</sup> |
| --- | --- | --- | --- | --- | --- |
| N (%) | 88 (32.5%) | 183 (67.5%) | 271 (100%) |  |  |
| **Demographics** |  |  |  |  |  |
| Age in years, mean (SD) | 17.83 (1.74) | 18.23 (2.07) | 18.10 (1.97) | -0.40 (0.24) | 0.09 |
| Female Gender, N (%) | 56 (63.6%) | 100 (54.6%) | 156 (57.6%) | 9.0% (6.3%) | 0.16 |
| Study-site = Porto Alegre | 38 (43.2%) | 70 (38.3%) | 108 (39.9%) | 4.9% (6.4%) | 0.44 |
| Low-risk demarcation <sup>a</sup> | 28 (31.8%) | 74 (40.4%) | 102 (37.6%) | -7.6% (6.1%) | 0.17 |
| **EMA Variables, mean (SD)** |  |  |  |  |  |
| Sadness <sup>b</sup> | 3.12 (1.32) | 3.40 (1.18) | 3.31 (1.23) | -0.28 (0.17) | 0.09 |
| Anxiety <sup>b</sup> | 3.41 (1.41) | 3.54 (1.32) | 3.50 (1.35) | -0.13 (0.18) | 0.44 |
| Inactivity <sup>b</sup> | 3.47 (1.32) | 3.77 (1.33) | 3.67 (1.33) | -0.30 (0.17) | 0.08 |
| Tiredness <sup>b</sup> | 4.07 (1.45) | 4.14 (1.30) | 4.12 (1.35) | -0.07 (0.18) | 0.70 |
| Irritability <sup>b</sup> | 3.13 (1.36) | 3.27 (1.31) | 3.22 (1.33) | -0.14 (0.17) | 0.42 |
| Overall mood <sup>b</sup> | 3.09 (1.30) | 3.18 (1.28) | 3.15 (1.29) | -0.09 (0.17) | 0.59 |
| Sleep quality <sup>c</sup> | 3.44 (1.19) | 3.24 (0.99) | 3.31 (1.06) | 0.20 (0.15) | 0.17 |

<sup>a</sup> The cohort was demarcated into a high-risk vs. low risk group based on based on high levels of symptoms within the family assessed by the family liability index.
<sup>b</sup> EMA variables covering sadness, anxiety, inactivity, tiredness, irritability, and overall mood are measured on a 1 to 7 Likert scale, where higher values correspond to poorer affect.
<sup>c</sup> Sleep quality is measured on a 1-5 Likert scale with higher values corresponding to better subjective sleep quality.
<sup>d</sup> Standard deviation of the difference is estimated using a Gaussian propagation of uncertainties.
<sup>e</sup> For continuous and Likert variables we used a two-sided t-test. Whereas for binary variables we used the two-proportion z-test.

### 3.2. Estimation of effects

Improved sleep quality decreased all affective symptoms as shown in Table 2. The greatest effect was on tiredness (ATE, −0.33, SD, 0.06, 95% equal-tailed credible interval (CI) [−0.45, −0.20]). The only effect that didn’t reach the two-sigma threshold was sleep quality on irritability (ATE, −0.09, SD, 0.06, 95% CI [−0.21, 0.03]). By contrast, improved affective symptoms had a smaller effect on that night’s sleep quality. The greatest effects were from anxiety (ATE, −0.11, SD, 0.05, 95% CI [−0.30, −0.02]) which was still approximately half the effect of sleep quality’s influence on anxiety (ATE, −0.23, SD, 0.05, 95% CI [−0.34, −0.13]).

**Table 2.** Estimated next-day effects between affective symptoms and sleep quality. We highlight levels of credible evidence at the two-sigma (†) and three-sigma (††) level.

|  |  | Mean | SD | Median | Q2.5 | Q97.5 | Level of Credible Evidence |
| --- | --- | --- | --- | --- | --- | --- | --- |
| Sleep Quality on Affective Symptoms | Anxiety | -0.23 | 0.05 | -0.24 | -0.34 | -0.13 | †† |
|  | Irritability | -0.09 | 0.06 | -0.10 | -0.21 | 0.03 |  |
|  | Inactivity | -0.22 | 0.05 | -0.22 | -0.32 | -0.12 | †† |
|  | Sadness | -0.20 | 0.05 | -0.20 | -0.31 | -0.10 | †† |
|  | Tiredness | -0.33 | 0.06 | -0.32 | -0.45 | -0.20 | †† |
|  | Overall mood | -0.21 | 0.05 | -0.21 | -0.30 | -0.12 | †† |
| Affective Symptoms on Sleep Quality | Anxiety | -0.11 | 0.05 | -0.11 | -0.21 | -0.02 | † |
|  | Irritability | -0.12 | 0.05 | -0.12 | -0.22 | -0.01 | † |
|  | Inactivity | -0.04 | 0.05 | -0.04 | -0.15 | 0.06 |  |
|  | Sadness | -0.03 | 0.05 | -0.03 | -0.14 | 0.07 |  |
|  | Tiredness | 0.01 | 0.05 | 0.01 | -0.09 | 0.10 |  |
|  | Overall mood | -0.08 | 0.06 | -0.07 | -0.19 | 0.03 |  |
| Lagged Sleep Quality on Affective Symptoms | Anxiety | -0.13 | 0.06 | -0.13 | -0.26 | -0.01 | † |
|  | Irritability | -0.12 | 0.05 | -0.12 | -0.23 | -0.02 | † |
|  | Inactivity | -0.10 | 0.06 | -0.10 | -0.22 | 0.01 |  |
|  | Sadness | -0.14 | 0.06 | -0.13 | -0.25 | -0.02 | † |
|  | Tiredness | -0.17 | 0.06 | -0.17 | -0.29 | -0.06 | † |
|  | Overall mood | -0.09 | 0.05 | -0.09 | -0.20 | 0.00 | † |

Improvements in lagged sleep quality also led to improvement on all affective variables after accounting for the prior night’s sleep quality. Although, typically at lower levels and meeting lower credible evidence thresholds. For example, the influence of prior night’s sleep quality on today’s anxiety (ATE, −0.13, SD, 0.06, 95% CI [−0.21, −0.01]), was about half that of the effect of last night’s sleep quality. This suggests that improvement in lagged sleep quality provides incremental improvement on affective symptoms.

We provide a broad summary of these findings in Figure 1. For more information regarding the relevant regression analyses, the fixed effects are summarised in Supplementary Material Tables S4-S6. We also show that there is no credible evidence for an interaction between high-risk selection and affective symptoms on sleep quality and vice versa in Supplementary Material 7.

**Figure 1.**
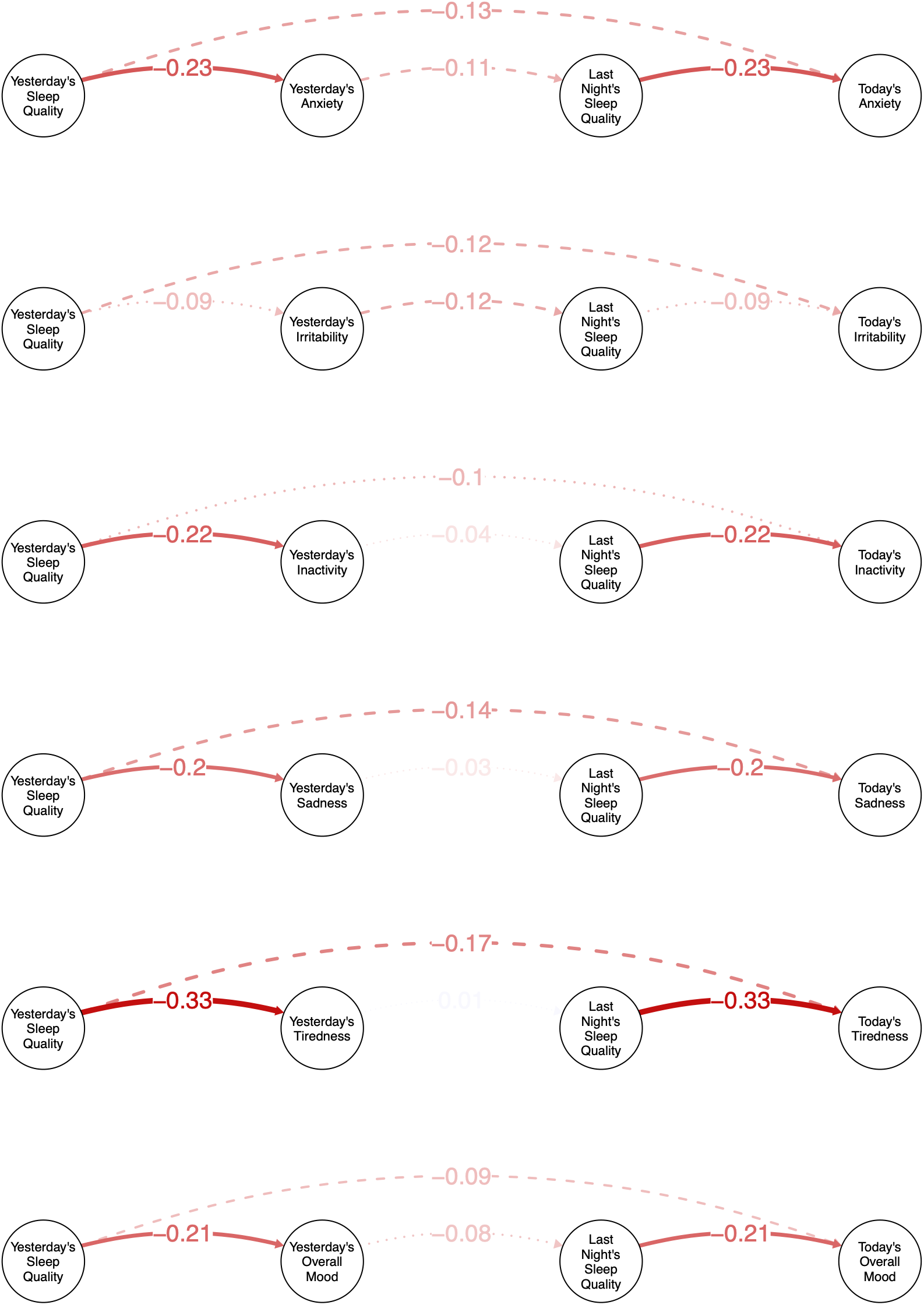
A graphical representation of the total effects estimated in our initial analysis. Consecutive night’s sleep quality has greater effects on affective symptoms than affective symptoms have on sleep quality. The expected average treatment effect (ATE) is represented quantitatively. We represent uncertainty at the three-sigma (solid), two-sigma (dashed), and less than two-sigma (dotted) levels.

We then estimated the effects of the four simulated consecutive day scenarios with comparisons between scenarios shown in Table 3. We found that the ‘stable good sleep’ scenario would improve all six affective symptoms compared to the ‘stable poor sleep’ scenario at the two-sigma level. Compared to ‘stable poor nights sleep’, the greatest effect of ‘stable good sleep’ was on overall mood (Δ, −2.10, SE, 1.03) whereas the weakest effect was on irritability (Δ, −1.21, SE, 0.67). Effects of the two mixed sleep quality scenarios corresponded to an approximate mid-point on affective symptoms between the stable good and poor sleep scenarios, although within the two-sigma level. Finally, the ‘worsening sleep’ scenario tended to outperform the ‘improving sleep’ scenario, although, all within the two-sigma level.

**Table 3.**
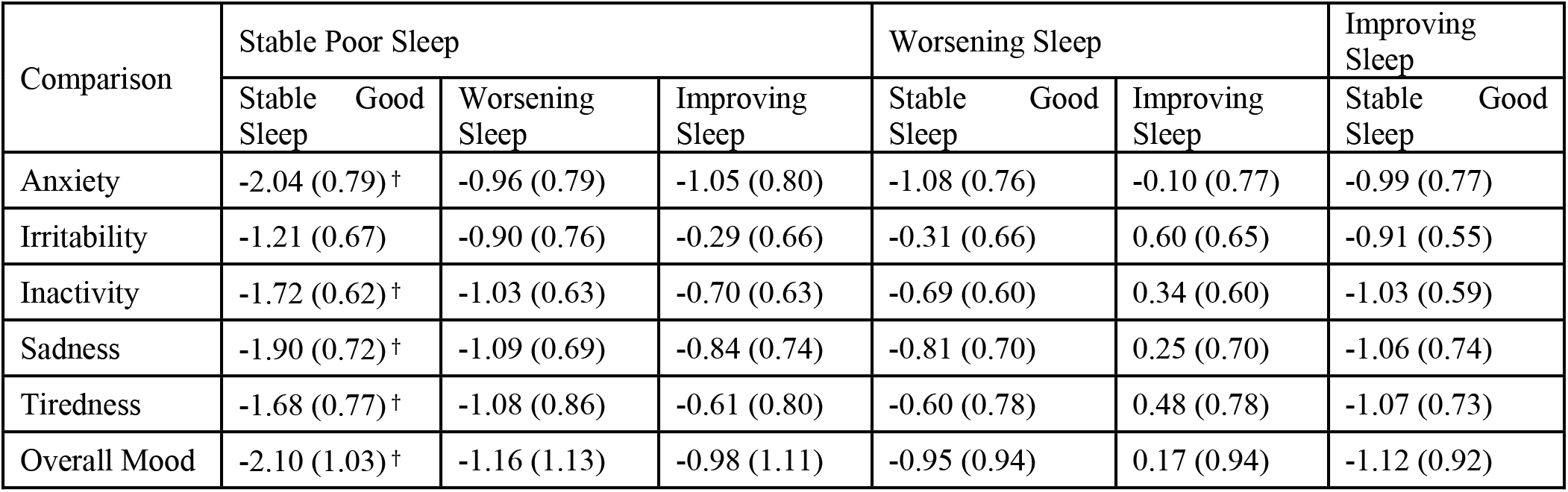
Comparison of expected affective symptoms under two-night sleep quality scenarios. We report the difference and standard error in expected affective symptoms under the scenario on the bottom row subtracted by the top row. We highlight levels of credible evidence at the two-sigma (†) and three-sigma (††) level.

## 4. Discussion

We found evidence for a within-person bidirectional feedback loops between day-to-day subjective sleep quality and self-rated affective symptoms, which is consistent with prior work (Hickman et al., 2024). Notably, sleep quality exerted greater influence on affective symptoms than the reverse. Sleep quality influenced all six affective symptoms, whereas affective symptoms had weaker effects on sleep quality, with only anxiety and irritability reaching the two-sigma threshold for credible evidence.

A novel contribution of this work is the estimation of the effect of changes in sleep quality across consecutive days influence affective symptoms. We showed this through two complementary methods. First, we showed that lagged sleep quality had an incremental effect on affective symptoms, even after adjusting for the prior night’s sleep quality, indicating that poor sleep has a cumulative, consecutive-day impact rather than a purely next-day effect. Second, we showed that improvements in sleep quality on consecutive days provided incremental improvements on affective symptoms. This is broadly consistent with previous results suggesting that full recovery from poor sleep quality took multiple days (Dinges et al., 1997), and with analyses using sleep duration in experimental sleep restriction settings (Booth et al., 2021) and naturalistic settings (Lee, 2022).

Our results advance our understanding of consecutive day effects by identifying and quantifying a dynamic mechanism between sleep and affect. Specifically, our simulations show that improvements in yesterday’s sleep quality improves lagged affective symptoms, which subsequently improves current day affective symptoms. Furthermore, our sequential scenarios isolate this mechanism by interrupting the path from affective symptoms to sleep quality, thereby illustrating how breaking the feedback loop can lead to incremental improvements in affective symptoms.

These findings provide mechanistic and temporal evidence consistent with a causal role for sleep-circadian disruption in the maintenance and progression of affective symptoms (Hickie and Crouse, 2024). We show that yesterday’s sleep quality exerts an independent effect on subsequent affective symptoms, including mood, and that this influence propagates through a dynamic feedback loop linking sleep and affect across days. Importantly, sleep quality from yesterday nights continued to influence affective symptoms over multiple subsequent days, even after adjustment for prior night’s sleep, indicating a temporally extended impact rather than a transient next-day effect. This consecutive-day propagation mirrors known circadian dynamics and supports a causal interpretation in which sleep disturbance functions as a slowly evolving control parameter that shapes affective trajectories over time, rather than merely reflecting downstream consequences of mood disorders.

### 4.1 Potential implication for treatments

Our analysis suggests that sustained improvements in sleep quality may be an effective treatment target for improving daily affective symptoms. This is consistent with prior literature that has suggested sleep quality may be an important intervention target for improving mood (Werner et al., 2025). Other research has highlighted the potential causal role of sleep disturbances on multiple psychiatric disorders (Freeman et al., 2020), suggesting that targeting sleep quality is likely to have broader clinical implications.

### 4.2. Potential drivers of consecutive poor sleep

Several mechanisms may underlie consecutive poor sleep quality with implications for treatment. The first is circadian dysregulation, which has been proposed as a key pathophysiological mechanism driving poor mood and mood disorders (Carpenter et al., 2021). Consecutive days of poor sleep quality may arise due to circadian misalignment where sleep behaviours are out of alignment with the internal body clock, or where circadian signals that play a role in sleep-wake regulation (e.g., cortisol, core body temperature) are disrupted *within* the circadian system (Carpenter et al., 2025; Chang et al., 2009). In these cases, therapies may need to more directly target a range of sleep-wake or circadian factors to improve sleep quality, and subsequently, affective symptoms. This may include behavioural, activity, social and work-related schedules, light-exposure, and potentially sleep and fatigue related pharmacological treatments (Crouse et al., 2021). Interventions correcting circadian disturbance are associated with improved mood (Wescott et al., 2025), and individuals with certain circadian features (which may reflect or manifest as disturbances) may be less responsive to non-circadian treatments such as selective serotonin reuptake inhibitors (Crouse et al., 2024). These effects are potentially through or independent of sleep quality, and thus they could also improve the overall cyclic regulatory system between affective symptoms and sleep quality. In non-clinical populations where inexpensive and low burden interventions may be particularly appropriate, increased exposure to daylight may be one public health recommendation that has the potential to positively impact sleep quality and affective symptoms (Crouse et al., 2025). Motor activity and subjective energy may also be important intervention targets for improving mood, as EMA evidence has highlighted these as having dynamic associations with sleep and mood (Merikangas et al., 2019).

Another potential mechanism that may underlie poor sleep quality is rumination. Rumination and sleep-wake disturbance have been identified as potential prognostic indicators of transdiagnostic transition to severe mental disorders (Grierson et al., 2019), and emerging evidence suggests that sleep quality may mediate the association between rumination (i.e., pre-sleep worrying) and next day affect (Werner et al., 2025). Consistent with this pathway, irritability, which is closely related to rumination, had the greatest effect on sleep quality in the current study. Together, these findings support a model in which rumination contributes to sleep disruption, which in turn amplifies affective symptoms, creating a self-reinforcing cycle. This positions rumination as a potentially modifiable target to improve mood and prevent mental ill health (Grierson et al., 2016).

### 4.3. Future Work

The analysis of sleep disturbances across consecutive days using observational studies is limited in the literature. We suspect that this is partially due to limited analytical approaches to perform such analyses. We adopted approaches from the causal inference literature and applied it to EMA data. We suggest that causal inference is an important approach to understand both next-day effects (e.g., Triantafillou et al., 2019) and consecutive-day effects as explored in our analysis. An appropriate next step would be to understand causal effects on longer-term sleep and affective symptom patterns. A particularly important consideration is that causal inference allows for a greater understanding of potential intervention targets and their effects (Prosperi et al., 2020; Sanchez et al., 2022), which could inform automated recommendations across interventions via comparison of outcomes under different scenarios (Varidel et al., 2024), allowing for potential real-time mental health recommendations.

Further, with the increasing collection and sharing of EMA data (Siepe et al., 2025), such analyses of observational and naturalistic data could be applied at larger scale and lower cost than intensive experimental designs. This could allow for greater understanding of causal mechanisms and estimation of effect sizes across different populations and conditions. At the same time, given that experimental designs remain the gold standard for causal inference, experimental designs could be informed by or combined with (Colnet et al., 2024) findings from wide-scale observational causal inference analyses.

Application of methods to identify individual-level causal mechanisms of sustained poor mental health outcomes could also find applications within digital mental health technologies (DMHTs), that frequently collect longitudinal data. Many existing methods to analyse such data emphasise day-to-day dynamics, which may not capture sustained poor mental health outcomes that such tools aim to improve. Methods that explicitly model sustained within-person effects, as applied here, may offer a more direct basis for personalising interventions.

### 4.4. Limitations

We only examined these sleep-affect relationships within a small Brazilian cohort with limited completion of the EMA protocol. Thus, our ability to probe to longer periods was limited. Further examination of these relationships in larger, more diverse, and longer-term samples will be important in understanding the generalisability of these findings. However, we do note that our findings are broadly consistent with other analyses (Booth et al., 2021; Hickman et al., 2024; Lee, 2022; Triantafillou et al., 2019), where comparisons can be made.

Subjective sleep quality provides insight into key aspects of an individual’s experience of sleep; however, this measure often does not reflect objective sleep parameters and is biased by mood itself (Windmill et al., 2024), which may partially explain the estimated bidirectional associations. It will be important for future work to examine both subjective and objective sleep measures simultaneously (e.g., portable polysomnography) to further understand these relationships. Further, future work should examine different aspects of subjective sleep quality (e.g. insomnia compared to non-restorative sleep) as these may be underpinned by different mechanistic processes and have different clinical implications and targets (Zhang et al., 2013). Finally, we did not examine within-day affective symptoms, instead averaging across observed within-day EMAs, which cannot fully capture the nuanced dynamic relationships between sleep quality and within-day variation in affective symptoms. Similarly, more objective measures of activity (e.g., step counts) could also be considered.

Methodologically, while our framework aims to infer causal effects, such interpretations must be tentative due to the observational nature of the study, primarily due to the potential for unobserved confounding. To combat this, we do account for key confounders including age, gender, high-risk cohort, and weekend day, which is consistent with prior approaches (e.g., Triantafillou et al., 2019). We also checked for biases between our sample and the underlying BHRC EMA cohort in the observed variables, to account for selection biases. Further, our analyses control for stable between-person differences by including random intercepts within our models. However, by controlling for stable between-person differences, which are often more significant than within-person differences, we may be missing important differences across the populations (Hamaker, 2023; Lüdtke and Robitzsch, 2021).

We also note that the EMA data was collected during the COVID-19 pandemic period. As such, sleep and affective patterns and their interactions may be different in other time periods. This moderately limits the generalisability of the study.

## 5. Conclusion

We found evidence for a within-person bidirectional feedback loop between sleep quality and affective symptoms, with sleep quality having greater influence on affective symptoms than vice versa. Novel to this work, we showed evidence that improvements in consecutive nights sleep quality had incremental improvements in all affective symptoms. Together, these results are consistent with affective symptoms and sleep quality operating through a feedback loop, with consecutive day improvements in sleep quality breaking that cycle. This supports the use of multifaceted and targeted treatments strategies that aim to improve sleep and circadian patterns over sustained periods.

## Credit authorship contribution statement

**Mathew Varidel:** Conceptualisation, Methodology, Writing – Original Draft, Writing – Review & Editing. **Luke Borgnolo:** Conceptualisation, Data curation, Writing – Original Draft. **Victor An:** Formal analysis. **Joanne Carpenter:** Writing – Review & Editing. **Ian Hickie:** Funding acquisition, Writing – Review & Editing. **Pedro M Pan:** Investigation, Writing – Review & Editing. **Francisco da Silva Jr**.: Data curation. **Jacob Crouse:** Funding acquisition, Writing – Review & Editing. **Eurípedes C Miguel:** Writing – Review & Editing. **Luís A Rohde:** Writing – Review & Editing. **Giovanni A Salum:** Writing – Review & Editing. **Frank Iorfino:** Conceptualisation, Supervision, Funding acquisition, Writing – Review & Editing.

## Data availability

Data was sourced from the Brazilian High-Risk Cohort Study (BHRC). Access to this data can be achieved by request from the relevant BHRC resources access through the project’s Open Science Framework page (https://osf.io/ktz5h/).

