## Supplementary Material for "Consecutive day effects between sleep quality and affective symptoms among youth in the Brazilian High-Risk Cohort study"

**Supplement 1. EMA Interface example**

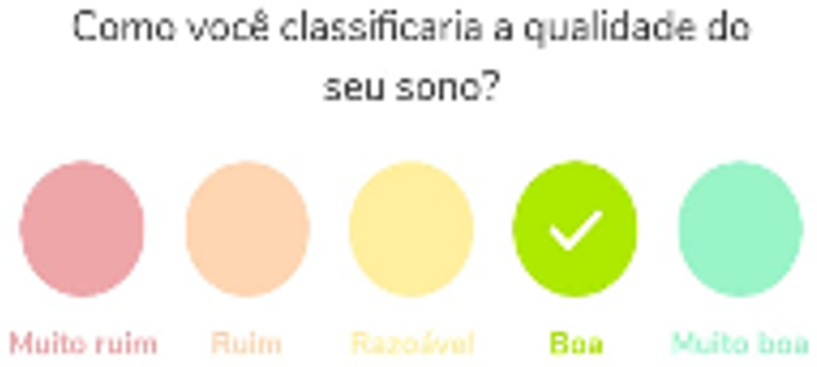


**Figure S1**. Sample question for the BHRC EMA question.

**Supplement 2.**

Here we provide more motivation and mathematical detail for the statistical analyses performed in this paper.

*Model Formulation*

We started by drawing a plausible structure representing the expected dependencies in Fig. S2. This structure can be broadly defined as a first-order vector autoregressive process. This structure is assumed throughout this manuscript to account for appropriate confounding variables.

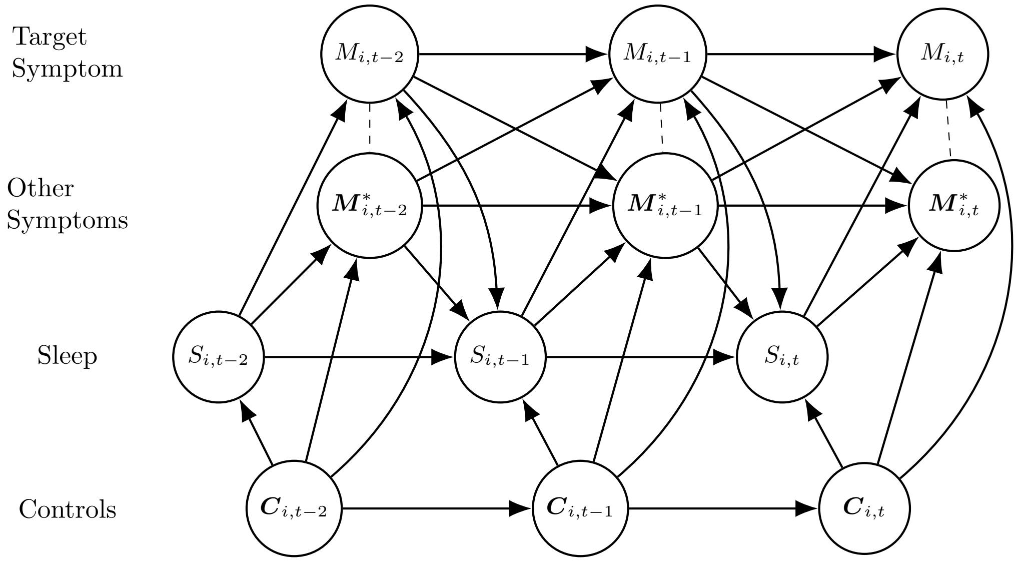


**Figure S2.** The assumed day-to-day within-person causal dependency structure. Throughout the article, we focus on total effects on a particular symptom for the current day denoted $M_{i,t}$ and sleep quality $S_{i,t}$ for an individual $i$. Therefore, to understand the total effect on the focus variable $M_{i,t}$, we consider covariation with other affective symptoms $\boldsymbol{M}^*_{i,t}$, as well as influence from last night's sleep quality $S_{i,t}$, dynamic controls $\boldsymbol{C}_{i,t}$, and prior day's affective symptoms $M_{i,t-1}$ and $\boldsymbol{M}^*_{i,t-1}$.

*Comparison of bidirectional effects between affective symptoms and sleep quality*

Consider the effect of prior night's sleep quality $S_{i,t}$ for individual $i$ on mood for the current day $t$ given by $M_{i,t}$. To provide an unbiased estimate of this the total within-person effect, we must select a set of controls that closes the backdoor paths to between $S_{i,t}$ and $M_{i,t}$ – that is we close paths that have paths that point to both variables (Pearl et al., 2016). Fig. S2 suggests that a total effect estimate of sleep quality on mood requires controlling for prior day's affective symptoms, $M_{i,t-1}$ and $\boldsymbol{M}^*_{i,t-1}$, dynamic controls $\boldsymbol{C}_{i,t} = \left(C_{i,t,1}, C_{i,t,2}, \ldots, C_{i,t,N_C}\right)$ using age, gender, study site, weekend day, and whether they were in the high-risk cohort. We estimate a mixed-effects linear regression model with these assumptions, defined as

$$M_{i,t} = \mu_i + (\mathrm{A} + \alpha_i)S_{i,t} + \Delta M_{i,t-1} + \sum_{j=1}^{N_{M^*}} \Phi_j^* M^*_{i,j,t-1} + \sum_{j=1}^{N_C} \kappa_j C_{i,t,j} + \epsilon_{i,t}.$$

The population-level total effect is represented by $A$ and $\alpha_{i,t}$ is individual $i$'s deviation from $A$. Then $\mu_i$ represents individual $i$'s time-invariant individual-level average for the target affective symptom. The remaining variables relate prior affective symptoms and controls to the current day target symptom assuming fixed slopes. Lastly, $\epsilon_{i,t}$ is a white noise contribution.

Next, consider the estimation of the effect of an individual's prior day symptom $M_{i,t-1}$ on prior night's sleep $S_{i,t}$ given the structure in Fig. S2. We highlight the unknown directionality between intraday symptoms. Given full knowledge of a causal structure represented by a directed acyclic graph we would be able to correct for intraday variables (Hernán and Robins, 2020; Loh and Ren, 2023). However, full knowledge of the intraday causal structure between affect variables is unknown. Therefore, we correct for prior day's mood $M_{i,t-2}$ and $M^*_{i,k,t-2}$ for $k = 1, \dots, K$, which is a common guideline to estimate total effects in epidemiological studies where the causal structure is not well known (VanderWeele, 2019). Although, we acknowledge that this approximation will likely lead to an over-estimation of the true total effect, as within-day influence is ignored. Once again, we assume a mixed effects linear regression model,

$$S_{i,t} = \nu_i + (B + \beta_i)M_{i,t-1} + \Gamma S_{i,t-1} + \Theta M_{i,t-2} + \sum_{j=1}^{N_{M^*}} \Theta^*_j M^*_{i,j,t-2} + \sum_{j=1}^{N_C} \lambda_j C_{i,t,j} + \zeta_{i,t}.$$

Where we follow similar notation to the above, such that $B$ represents the population-level total effect and $\beta_{i,t}$ represents individual $i$'s deviation of their total effect. Then $\nu_i$ represents an individual's average sleep quality and $\zeta_{i,t}$ is a white noise contribution. The remaining parameters are used as adjustment factors, which for simplicity we assume fixed slopes for each parameter.

We note that a common alternative is to estimate the direct effect of $M_{i,t-1}$ on $S_{i,t}$ by controlling for all other lagged affective symptoms $\boldsymbol{M}^*_{i,t-1}$. However, we wish to compare the total effect of $S_{i,t}$ on $M_{i,t}$ to the total effect of $M_{i,t-1}$ on $S_{i,t}$. Estimating the direct effect will underestimate this total effect and thus make the comparison inappropriate.

*Influence of previous night's sleep quality on affective symptoms*

We then targeted the total effect from lagged sleep quality $S_{i,t-1}$ independent from last night's sleep $S_{i,t}$. First, we remove the total effect of prior night's sleep on affective symptoms following Loh and Ren (2023),

$$R_{i,t,-S_t} = M_{i,t} - (\widehat{\mathrm{A}} + \hat{\alpha}_i)S_{i,t}.$$

The hatted symbols denote estimated parameters. Then, we estimate a mixed-effects linear regression model on $R_{i,t,-S_t}$ due to lagged sleep and accounting for confounders as identified under the assumed structure defined in Fig. S2., such that

$$R_{i,t,-S_t} = \lambda_i + (\Omega + \omega_i)S_{i,t-1} + Z M_{i,t-2} + \sum_{j=1}^{N_{M^*}} \Psi^*_j M^*_{i,t-2} + \sum_{j=1}^{N_C} \pi_j C_{i,t,j} + \tau_{i,t}.$$

*Influence of consecutive nights sleep quality on affective symptoms*

We then investigated the effect of sequential interventions on sleep quality. Broadly following g-estimation procedures (Hernán and Robins, 2020), we target the causal effect between two different deterministic sequential intervention strategies. That is, we propose a sequential treatment that deterministically sets lagged sleep quality to $s^*_{t-1}$ and prior night's sleep quality to $s^*_t$ for all individuals denoted by $do(S_{i,t-1} = s^*_{t-1}, S_{i,t} = s^*_t)$. Fig. S3 provides a schematic for a hypothetical scenario. We then aim to estimate the expected value under this scenario given by $E\left[M_{i,t}^{S_{i,t-1}=s^*_{t-1}, S_{i,t}=s^*_t}\right]$. To estimate this we integrate (i.e., average over) the intermediate mood variables $M_{i,t-1}, M^*_{i,1,t-1}, \dots, M^*_{i,N_{M^*},t-1}$ given the intervention $S_{i,t-1} = s^*_{t-1}$. More precisely, we evaluate,

$$E\left[M_{i,t}^{S_{i,t-1}=s^*_{t-1}, S_{i,t}=s^*_t}\right] = \iint dM_{i,t-1} d\boldsymbol{M}^*_{i,1,t-1} E\left[M_{i,t} | S_{i,t} = s^*_t, M_{i,t-1}, \boldsymbol{M}^*_{i,t-1}, \boldsymbol{C}_i\right] p(M_{i,t-1}, \boldsymbol{M}^*_{i,t-1} | S_{i,t-1} = s^*_{t-1}, M_{i,t-2}, \boldsymbol{M}^*_{i,1,t-2}, \boldsymbol{C}_i).$$

Therefore, we estimate the joint distribution model,

$$\begin{pmatrix} M_{i,t-1} \\ M^*_{i,1,t-1} \\ \cdots \\ M^*_{i,N_{M^*},t-1} \end{pmatrix} = \boldsymbol{\mu}_i + \left(S_{i,t-1}, M_{i,t-2}, M^*_{i,1,t-2}, \dots, M^*_{i,N_{M^*},t-2}, C_{i,1}, \dots, C_{i,N_C}\right)\underline{D} + \boldsymbol{\epsilon}_{i,t}$$

Where $\underline{\boldsymbol{D}}_i$ is a $(2 + N_{M^*} + N_C) \times (2 + N_{M^*} + N_C)$ is real-valued transition matrix, following the previous approach we estimate random effects for lagged sleep on mood and fixed effects otherwise. Then $\boldsymbol{\epsilon}_{i,t}$ is multivariate noise estimated as a fixed-effects covariance matrix. We chose to integrate this through simulation. This joint distribution, along with the regression model for the next day is then used to simulate an affective symptom for the following day. We then average over the individual-level samples of simulated affective symptoms. Comparison of two hypothetical sequential scenarios is estimated as the difference in expectation for each scenario averaged across the population,

$$\Delta_{*\dagger} = E\left[M^{S_{t-1}=s_{t-1}^*, S_t=s_t^*}\right] - E\left[M^{S_{t-1}=s_{t-1}^\dagger, S_t=s_t^\dagger}\right].$$

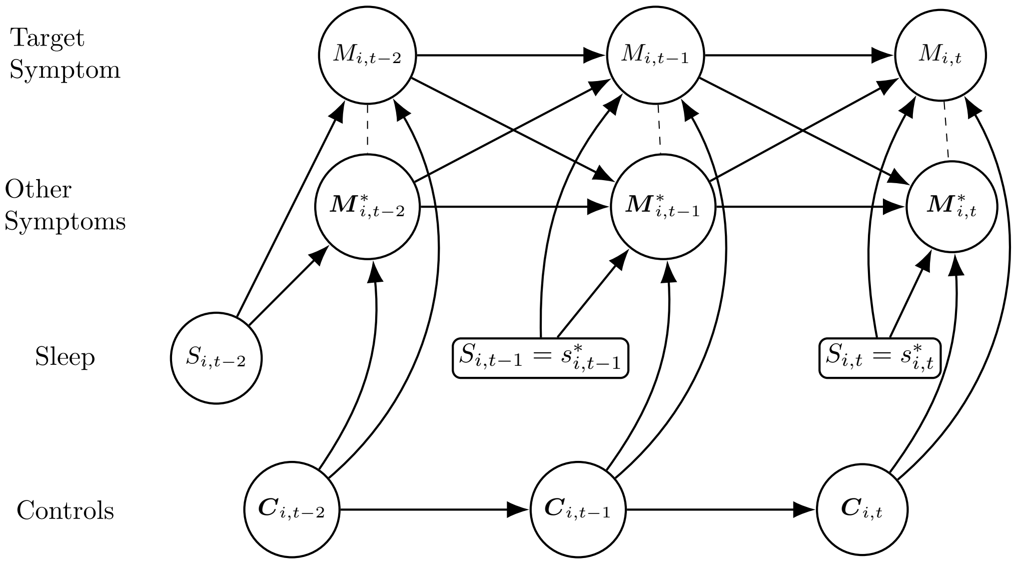


**Figure S3.** The assumed within-person causal dependency structure under hypothetical interventions on lagged ($S_{i,t-2}$) and last night's ($S_{i,t-1}$) sleep quality for individual $i$. The consecutive day interventions act on affective symptoms by directly influencing next-day affective symptoms and by breaking the feedback loops between lagged sleep quality and affective symptoms on last night's sleep quality.

**Supplement 3.** Flowchart for selection criteria for each analysis.

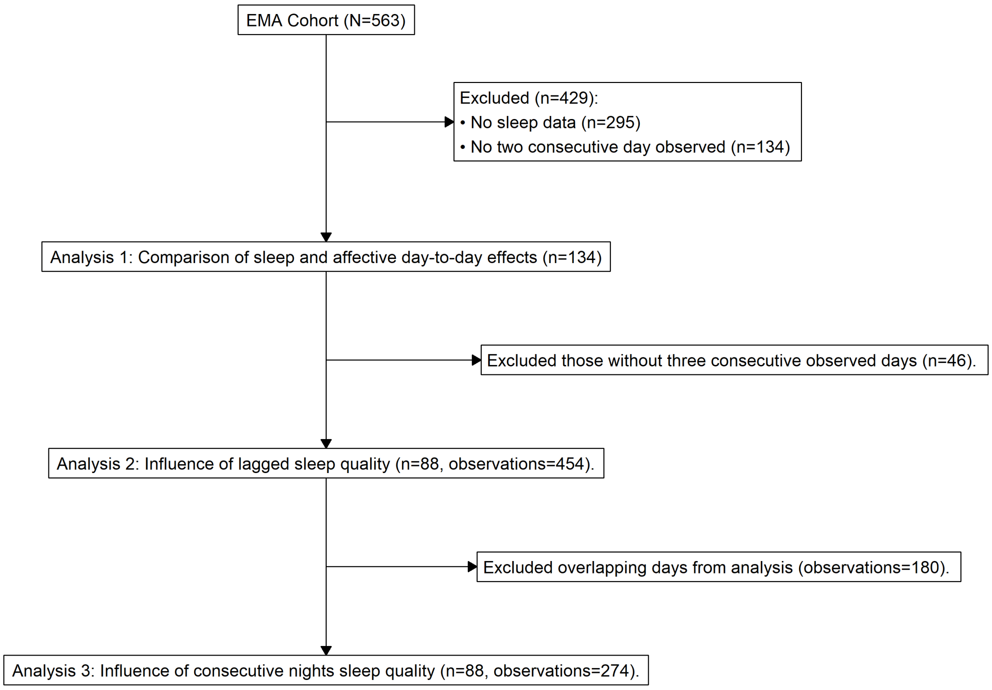


**Figure S4.** Selection criteria for each analysis.

**Supplement 4. Summary of fixed effects when targeting sleep quality on affective symptoms**

**Table S1.** Fixed effect regression coefficients when targeting sleep quality on Anxiety.

| | **Mean** | **SD** | **Q2.5** | **Median** | **Q97.5** | **N_eff** | **Rhat** |
|---|---|---|---|---|---|---|---|
| (Intercept) | -0.6773 | 0.517139 | -1.72674 | -0.66837 | 0.322069 | 8775 | 0.999803 |
| sleep_quality | -0.23476 | 0.054067 | -0.33939 | -0.23535 | -0.12765 | 10580 | 0.999901 |
| lag_sadness | -0.04808 | 0.101561 | -0.25281 | -0.04734 | 0.148874 | 7760 | 1.000477 |
| lag_anxiety | 0.227414 | 0.061704 | 0.107627 | 0.226837 | 0.350275 | 9920 | 0.999992 |
| lag_inactivity | -0.05342 | 0.081323 | -0.21168 | -0.05357 | 0.103899 | 9395 | 1.000336 |
| lag_tiredness | 0.103383 | 0.062472 | -0.01787 | 0.102918 | 0.227257 | 10303 | 1.000179 |
| lag_irritability | 0.224362 | 0.073773 | 0.08001 | 0.224922 | 0.366367 | 12345 | 0.999744 |
| lag_overall_mood | 0.072961 | 0.089715 | -0.09982 | 0.072908 | 0.2487 | 8959 | 1.000003 |
| lag_sleep_quality | 0.011706 | 0.054474 | -0.09573 | 0.011916 | 0.117874 | 11685 | 0.999961 |
| site | 0.028534 | 0.069795 | -0.1081 | 0.027835 | 0.166039 | 12714 | 0.999656 |
| gender | 0.014776 | 0.075093 | -0.13317 | 0.015334 | 0.163925 | 10712 | 0.999763 |
| age | 0.029611 | 0.021524 | -0.01203 | 0.029158 | 0.072323 | 10014 | 0.999639 |
| selection | 0.059702 | 0.076936 | -0.09241 | 0.059825 | 0.211732 | 10991 | 0.999983 |
| is_weekend | -0.03222 | 0.070463 | -0.17281 | -0.03181 | 0.105689 | 13219 | 0.999897 |

**Table S2.** Fixed effect regression coefficients when targeting sleep quality on irritability.

| | **Mean** | **SD** | **Q2.5** | **Median** | **Q97.5** | **N_eff** | **Rhat** |
|---|---|---|---|---|---|---|---|
| (Intercept) | 0.383833 | 0.473134 | -0.54928 | 0.383842 | 1.327853 | 6296 | 1.00123 |
| sleep_quality | -0.09482 | 0.059042 | -0.20519 | -0.09622 | 0.027674 | 5940 | 1.000403 |
| lag_sadness | 0.235631 | 0.09152 | 0.058697 | 0.234832 | 0.418813 | 6974 | 1.000369 |
| lag_anxiety | 0.00368 | 0.055615 | -0.10667 | 0.00388 | 0.111125 | 7983 | 1.000176 |
| lag_inactivity | -0.07728 | 0.074739 | -0.22432 | -0.07734 | 0.068544 | 7882 | 1.001048 |
| lag_tiredness | 0.093243 | 0.057828 | -0.01873 | 0.093516 | 0.208049 | 9045 | 1.000611 |
| lag_irritability | 0.330957 | 0.06496 | 0.203491 | 0.33085 | 0.458112 | 7907 | 1.000274 |
| lag_overall_mood | -0.07174 | 0.080342 | -0.22977 | -0.0721 | 0.085589 | 7861 | 0.999729 |
| lag_sleep_quality | 0.020177 | 0.049493 | -0.07654 | 0.020216 | 0.119101 | 8085 | 0.999837 |
| site | 0.018175 | 0.065491 | -0.10966 | 0.018113 | 0.145865 | 9536 | 0.999606 |
| gender | -0.05786 | 0.07072 | -0.19555 | -0.05762 | 0.081903 | 5020 | 1.00114 |
| age | -0.01973 | 0.019505 | -0.05781 | -0.01956 | 0.018263 | 8671 | 1.000855 |
| selection | 0.014129 | 0.070646 | -0.12157 | 0.013713 | 0.152411 | 7722 | 1.000431 |
| is_weekend | 0.00277 | 0.062468 | -0.11998 | 0.003124 | 0.125693 | 11088 | 0.999867 |

**Table S3.** Fixed effect regression coefficients when targeting sleep quality on overall mood.

| | **Mean** | **SD** | **Q2.5** | **Median** | **Q97.5** | **N_eff** | **Rhat** |
|---|---|---|---|---|---|---|---|
| (Intercept) | -0.3557 | 0.445423 | -1.23976 | -0.35721 | 0.509768 | 11679 | 0.999777 |
| sleep_quality | -0.20736 | 0.045394 | -0.29555 | -0.20772 | -0.1177 | 14203 | 0.999628 |
| lag_sadness | 0.120036 | 0.087783 | -0.05313 | 0.121743 | 0.293132 | 11322 | 0.999834 |
| lag_anxiety | -0.01241 | 0.052757 | -0.11488 | -0.01203 | 0.09244 | 15057 | 0.999684 |
| lag_inactivity | -0.0551 | 0.073556 | -0.19942 | -0.05515 | 0.086992 | 13861 | 0.999738 |

| | | | | | | | |
|---|---|---|---|---|---|---|---|
| lag_tiredness | 0.106711 | 0.056209 | -0.00138 | 0.105954 | 0.218732 | 15651 | 0.999662 |
| lag_irritability | 0.111745 | 0.06265 | -0.01275 | 0.111018 | 0.234546 | 17497 | 0.999762 |
| lag_overall_mood | 0.265091 | 0.078536 | 0.11075 | 0.264829 | 0.418631 | 14093 | 0.999631 |
| lag_sleep_quality | 0.088914 | 0.047883 | -0.00566 | 0.089482 | 0.181176 | 16954 | 0.999651 |
| site | -0.03788 | 0.060887 | -0.15807 | -0.03758 | 0.079667 | 16837 | 0.999664 |
| gender | 0.001147 | 0.065266 | -0.12498 | 0.000884 | 0.128833 | 13961 | 0.999753 |
| age | 0.016876 | 0.018296 | -0.01922 | 0.017388 | 0.052937 | 12432 | 0.999836 |
| selection | 0.073853 | 0.066254 | -0.05665 | 0.073554 | 0.205895 | 14006 | 0.999719 |
| is_weekend | 0.028255 | 0.060834 | -0.09093 | 0.027552 | 0.149098 | 18205 | 0.999661 |

**Table S4.** Fixed effect regression coefficients when targeting sleep quality on inactivity.

| | **Mean** | **SD** | **Q2.5** | **Median** | **Q97.5** | **N_eff** | **Rhat** |
|---|---|---|---|---|---|---|---|
| (Intercept) | 0.559177 | 0.511952 | -0.43431 | 0.558033 | 1.566954 | 14735 | 0.999851 |
| sleep_quality | -0.22375 | 0.051498 | -0.32407 | -0.22411 | -0.12387 | 17783 | 0.999656 |
| lag_sadness | 0.117049 | 0.100363 | -0.07803 | 0.11673 | 0.315192 | 14306 | 0.999766 |
| lag_anxiety | -0.1287 | 0.061297 | -0.24714 | -0.12846 | -0.01037 | 16021 | 0.99969 |
| lag_inactivity | 0.238413 | 0.082823 | 0.075256 | 0.238599 | 0.40224 | 15542 | 0.999884 |
| lag_tiredness | 0.203322 | 0.06343 | 0.078419 | 0.203099 | 0.32627 | 17856 | 0.999671 |
| lag_irritability | 0.117344 | 0.07017 | -0.02047 | 0.11683 | 0.253387 | 18162 | 0.999585 |
| lag_overall_mood | -0.04417 | 0.091413 | -0.224 | -0.04438 | 0.138009 | 16589 | 0.999771 |
| lag_sleep_quality | 0.065938 | 0.054516 | -0.04206 | 0.065945 | 0.172292 | 21325 | 0.999571 |
| site | -0.0555 | 0.06864 | -0.18788 | -0.05562 | 0.076577 | 16009 | 0.999717 |
| gender | -0.09277 | 0.075457 | -0.24262 | -0.09278 | 0.052522 | 16908 | 0.999824 |
| age | -0.02005 | 0.021078 | -0.06179 | -0.01987 | 0.020978 | 15121 | 0.999842 |
| selection | 0.012473 | 0.074348 | -0.13283 | 0.01243 | 0.155981 | 15455 | 0.999748 |
| is_weekend | 0.049707 | 0.068941 | -0.08596 | 0.049694 | 0.186424 | 19023 | 0.999638 |

**Table S5.** Fixed effect regression coefficients when targeting sleep quality on sadness.

| | **Mean** | **SD** | **Q2.5** | **Median** | **Q97.5** | **N_eff** | **Rhat** |
|---|---|---|---|---|---|---|---|
| (Intercept) | 0.113998 | 0.489963 | -0.85432 | 0.106461 | 1.067569 | 7563 | 1.000065 |
| sleep_quality | -0.2025 | 0.054274 | -0.30713 | -0.20345 | -0.09538 | 8608 | 0.999911 |
| lag_sadness | 0.218914 | 0.095052 | 0.029746 | 0.219298 | 0.403864 | 7190 | 0.999881 |
| lag_anxiety | -0.02332 | 0.057849 | -0.13488 | -0.02393 | 0.090966 | 10950 | 0.999947 |
| lag_inactivity | 0.0272 | 0.079096 | -0.12334 | 0.02734 | 0.182213 | 8367 | 0.999675 |
| lag_tiredness | 0.138034 | 0.059929 | 0.019338 | 0.138509 | 0.254806 | 9793 | 0.999826 |
| lag_irritability | 0.101673 | 0.068704 | -0.03331 | 0.101352 | 0.236008 | 10650 | 0.999871 |
| lag_overall_mood | 0.084141 | 0.08607 | -0.08472 | 0.084807 | 0.251033 | 7950 | 0.999704 |
| lag_sleep_quality | 0.046775 | 0.05152 | -0.05523 | 0.047195 | 0.14847 | 11959 | 0.99962 |
| site | -0.06221 | 0.06613 | -0.19189 | -0.0629 | 0.068124 | 7566 | 1.00019 |
| gender | -0.06048 | 0.070463 | -0.19596 | -0.06166 | 0.07944 | 9843 | 0.999989 |
| age | 0.002699 | 0.020216 | -0.03696 | 0.002777 | 0.042015 | 9500 | 0.999875 |
| selection | 0.021411 | 0.073595 | -0.12399 | 0.021435 | 0.164349 | 10504 | 0.99979 |
| is_weekend | 0.060157 | 0.066613 | -0.06955 | 0.060348 | 0.190796 | 12880 | 0.999868 |

**Table S6.** Fixed effect regression coefficients when targeting sleep quality on tiredness.

| | Mean | SD | Q2.5 | Median | Q97.5 | N_eff | Rhat |
|---|---|---|---|---|---|---|---|
| (Intercept) | 0.665236 | 0.561996 | -0.44788 | 0.665617 | 1.767164 | 7933 | 1.000594 |
| sleep_quality | -0.32629 | 0.06391 | -0.45236 | -0.32494 | -0.20297 | 8752 | 1.000197 |
| lag_sadness | 0.172423 | 0.104396 | -0.03224 | 0.173498 | 0.373668 | 8173 | 1.000302 |
| lag_anxiety | -0.08846 | 0.063725 | -0.2146 | -0.08929 | 0.037792 | 9541 | 1.000087 |
| lag_inactivity | 0.116059 | 0.085774 | -0.05205 | 0.116192 | 0.284825 | 8949 | 1.001076 |
| lag_tiredness | 0.252666 | 0.06572 | 0.122447 | 0.253521 | 0.379394 | 10280 | 1.000382 |
| lag_irritability | 0.052931 | 0.073336 | -0.08875 | 0.051917 | 0.196727 | 11049 | 0.999921 |
| lag_overall_mood | -0.09658 | 0.091149 | -0.27287 | -0.09686 | 0.082568 | 10225 | 0.999801 |
| lag_sleep_quality | -0.0195 | 0.056005 | -0.12908 | -0.02002 | 0.090278 | 12831 | 0.99964 |
| site | -0.04193 | 0.076009 | -0.18968 | -0.04147 | 0.108703 | 9730 | 0.999775 |
| gender | -0.0097 | 0.080418 | -0.16543 | -0.01118 | 0.146441 | 9530 | 1.000086 |
| age | -0.03031 | 0.023343 | -0.07611 | -0.03002 | 0.015535 | 9664 | 1.000224 |
| selection | -0.03617 | 0.082845 | -0.1964 | -0.03644 | 0.128291 | 8687 | 1.000449 |
| is_weekend | -0.06007 | 0.072175 | -0.19876 | -0.05989 | 0.081565 | 13307 | 0.999781 |

**Supplement 5. Summary of fixed effects when targeting affective symptom effects on sleep quality.**

**Table S7.** Fixed effect regression coefficients when targeting anxiety on sleep quality.

| | **Mean** | **SD** | **Q2.5** | **Median** | **Q97.5** | **N_eff** | **Rhat** |
|---|---|---|---|---|---|---|---|
| (Intercept) | -0.02032 | 0.486216 | -0.9554 | -0.02231 | 0.935363 | 13172 | 0.999867 |
| lag_anxiety | -0.11413 | 0.048131 | -0.2078 | -0.11414 | -0.02148 | 13397 | 0.999728 |
| lag_sleep_quality | -0.06757 | 0.051724 | -0.16885 | -0.0682 | 0.036283 | 14668 | 0.999858 |
| site | -0.02184 | 0.068553 | -0.15661 | -0.02162 | 0.114052 | 16872 | 1.000022 |
| gender | 0.039362 | 0.070331 | -0.09911 | 0.039775 | 0.179283 | 15620 | 0.999651 |
| age | 0.005613 | 0.020138 | -0.03365 | 0.005816 | 0.044705 | 13687 | 0.999989 |
| selection | -0.04144 | 0.072227 | -0.18386 | -0.04076 | 0.102483 | 13638 | 0.999852 |
| twice_lag_sadness | -0.14753 | 0.09389 | -0.33383 | -0.14632 | 0.035462 | 12662 | 0.999942 |
| twice_lag_anxiety | 0.053238 | 0.059686 | -0.06475 | 0.053267 | 0.169326 | 14549 | 1 |
| twice_lag_inactivity | 0.071381 | 0.076676 | -0.07627 | 0.071669 | 0.222144 | 12117 | 1.00012 |
| twice_lag_tiredness | 0.014209 | 0.059845 | -0.10354 | 0.01422 | 0.132157 | 14940 | 0.999779 |
| twice_lag_irritability | -0.13539 | 0.067708 | -0.26814 | -0.13529 | -0.00297 | 15283 | 0.999601 |
| twice_lag_overall_mood | 0.137931 | 0.077942 | -0.0152 | 0.138243 | 0.28939 | 16150 | 0.999706 |
| is_weekend | 0.033331 | 0.069205 | -0.09987 | 0.032688 | 0.168818 | 15889 | 0.999637 |
| is_lag_weekend | -0.0856 | 0.072304 | -0.22625 | -0.08573 | 0.057501 | 16584 | 0.999625 |

**Table S8.** Fixed effect regression coefficients when targeting irritability on sleep quality.

| | **Mean** | **SD** | **Q2.5** | **Median** | **Q97.5** | **N_eff** | **Rhat** |
|---|---|---|---|---|---|---|---|
| (Intercept) | 0.014615 | 0.48156 | -0.93284 | 0.014781 | 0.958703 | 13945 | 0.999675 |
| lag_irritability | -0.11598 | 0.053197 | -0.22067 | -0.11621 | -0.01298 | 14705 | 0.999597 |
| lag_sleep_quality | -0.06343 | 0.050563 | -0.16608 | -0.06318 | 0.034851 | 16754 | 0.999623 |
| site | -0.02042 | 0.066982 | -0.15176 | -0.01937 | 0.111244 | 14151 | 0.999588 |
| gender | 0.042682 | 0.070936 | -0.09589 | 0.041537 | 0.183091 | 13839 | 0.999706 |
| age | 0.004039 | 0.020238 | -0.03546 | 0.003999 | 0.044114 | 15027 | 0.99977 |
| selection | -0.05242 | 0.073678 | -0.19542 | -0.05267 | 0.090989 | 14549 | 0.999676 |
| twice_lag_sadness | -0.11325 | 0.097007 | -0.30271 | -0.1126 | 0.076333 | 11025 | 0.999727 |
| twice_lag_anxiety | 0.024105 | 0.058197 | -0.08921 | 0.024228 | 0.139051 | 14702 | 0.999651 |
| twice_lag_inactivity | 0.062893 | 0.076553 | -0.08784 | 0.062953 | 0.212265 | 13394 | 0.999641 |
| twice_lag_tiredness | 0.011424 | 0.05988 | -0.10589 | 0.012377 | 0.127701 | 16779 | 0.999753 |
| twice_lag_irritability | -0.11968 | 0.069384 | -0.25561 | -0.12013 | 0.016144 | 14867 | 0.999894 |
| twice_lag_overall_mood | 0.121025 | 0.079533 | -0.03777 | 0.122889 | 0.275037 | 14189 | 0.999874 |
| is_weekend | 0.040804 | 0.070218 | -0.09509 | 0.041024 | 0.177155 | 19674 | 0.999819 |
| is_lag_weekend | -0.07277 | 0.073378 | -0.21465 | -0.07245 | 0.06748 | 16446 | 0.999578 |

**Table S9.** Fixed effect regression coefficients when targeting overall mood on sleep quality.

| | **Mean** | **SD** | **Q2.5** | **Median** | **Q97.5** | **N_eff** | **Rhat** |
|---|---|---|---|---|---|---|---|
| (Intercept) | -0.02043 | 0.491688 | -0.98896 | -0.02521 | 0.933928 | 11348 | 0.999593 |
| lag_overall_mood | -0.07547 | 0.055994 | -0.18633 | -0.07449 | 0.034409 | 11548 | 1.000297 |
| lag_sleep_quality | -0.05983 | 0.051781 | -0.1623 | -0.05974 | 0.040492 | 13624 | 1.000181 |
| site | -0.01874 | 0.069313 | -0.15444 | -0.0196 | 0.115571 | 13948 | 0.999764 |
| gender | 0.042029 | 0.072226 | -0.0978 | 0.042401 | 0.185472 | 12085 | 0.999791 |
| age | 0.005013 | 0.020383 | -0.03576 | 0.005289 | 0.045052 | 12536 | 0.999528 |
| selection | -0.04301 | 0.074172 | -0.18885 | -0.04298 | 0.10081 | 12830 | 0.999767 |
| twice_lag_sadness | -0.12933 | 0.097393 | -0.32093 | -0.1302 | 0.060986 | 12247 | 0.999598 |
| twice_lag_anxiety | 0.020461 | 0.059132 | -0.09481 | 0.020486 | 0.139951 | 14572 | 0.999596 |
| twice_lag_inactivity | 0.075829 | 0.078802 | -0.07671 | 0.074979 | 0.231739 | 13083 | 0.999744 |
| twice_lag_tiredness | 0.009982 | 0.05963 | -0.10683 | 0.009509 | 0.126456 | 13222 | 0.999602 |
| twice_lag_irritability | -0.14468 | 0.068729 | -0.27815 | -0.14445 | -0.01088 | 13615 | 0.999658 |
| twice_lag_overall_mood | 0.154549 | 0.079566 | -0.00049 | 0.154575 | 0.31009 | 13058 | 0.999648 |
| is_weekend | 0.040788 | 0.069935 | -0.09806 | 0.04102 | 0.176942 | 14734 | 0.999766 |
| is_lag_weekend | -0.07935 | 0.072763 | -0.22303 | -0.07883 | 0.060637 | 13826 | 0.999794 |

**Table S10.** Fixed effect regression coefficients when targeting inactivity on sleep quality.

| | **Mean** | **SD** | **Q2.5** | **Median** | **Q97.5** | **N_eff** | **Rhat** |
|---|---|---|---|---|---|---|---|
| (Intercept) | 0.005281 | 0.490463 | -0.92781 | 0.001979 | 0.976283 | 7468 | 1.000313 |
| lag_inactivity | -0.04325 | 0.053269 | -0.14985 | -0.04265 | 0.059719 | 9745 | 0.999874 |
| lag_sleep_quality | -0.04965 | 0.052134 | -0.15141 | -0.05011 | 0.051979 | 11306 | 0.999825 |
| site | -0.02216 | 0.068951 | -0.15593 | -0.02254 | 0.112559 | 10246 | 0.999755 |
| gender | 0.041971 | 0.071073 | -0.10166 | 0.04192 | 0.182736 | 9459 | 1.000014 |
| age | 0.004236 | 0.020312 | -0.03577 | 0.004558 | 0.043484 | 8704 | 1.000222 |
| selection | -0.04803 | 0.073719 | -0.19291 | -0.0492 | 0.096783 | 10709 | 0.999603 |
| twice_lag_sadness | -0.13652 | 0.093063 | -0.32181 | -0.13677 | 0.044812 | 7161 | 1.000147 |
| twice_lag_anxiety | 0.015589 | 0.058887 | -0.10051 | 0.015068 | 0.128869 | 9921 | 1.000195 |
| twice_lag_inactivity | 0.088571 | 0.077341 | -0.06487 | 0.088661 | 0.242372 | 7961 | 0.999709 |
| twice_lag_tiredness | 0.012172 | 0.059422 | -0.10205 | 0.011929 | 0.129163 | 10799 | 1.000133 |
| twice_lag_irritability | -0.14424 | 0.067757 | -0.27843 | -0.14466 | -0.01048 | 10973 | 0.99969 |
| twice_lag_overall_mood | 0.135474 | 0.078956 | -0.0195 | 0.135418 | 0.29103 | 10335 | 0.999644 |
| is_weekend | 0.043685 | 0.069767 | -0.09287 | 0.043127 | 0.181829 | 13058 | 0.999674 |
| is_lag_weekend | -0.08537 | 0.072667 | -0.22889 | -0.08542 | 0.057806 | 12266 | 0.999719 |

**Table S11.** Fixed effect regression coefficients when targeting sadness on sleep quality.

| | **Mean** | **SD** | **Q2.5** | **Median** | **Q97.5** | **N_eff** | **Rhat** |
|---|---|---|---|---|---|---|---|
| (Intercept) | -0.03476 | 0.483144 | -0.99275 | -0.03416 | 0.901638 | 9712 | 1.00093 |
| lag_sadness | -0.03134 | 0.053303 | -0.13578 | -0.03174 | 0.072886 | 10336 | 0.999811 |
| lag_sleep_quality | -0.04538 | 0.050898 | -0.14487 | -0.04577 | 0.053472 | 12033 | 0.999746 |
| site | -0.0199 | 0.068427 | -0.15301 | -0.02028 | 0.1131 | 12564 | 0.999821 |
| gender | 0.044139 | 0.071512 | -0.09471 | 0.044887 | 0.182658 | 11639 | 0.999845 |
| age | 0.006035 | 0.020035 | -0.03345 | 0.006038 | 0.046434 | 11044 | 1.000663 |
| selection | -0.04499 | 0.073356 | -0.18575 | -0.04516 | 0.09887 | 12114 | 1.000442 |
| twice_lag_sadness | -0.13316 | 0.096633 | -0.32204 | -0.1335 | 0.05531 | 8234 | 0.999906 |
| twice_lag_anxiety | 0.020593 | 0.058975 | -0.09366 | 0.021512 | 0.135145 | 10731 | 1.000176 |
| twice_lag_inactivity | 0.079104 | 0.077987 | -0.0733 | 0.079404 | 0.229599 | 8644 | 0.999677 |
| twice_lag_tiredness | 0.00821 | 0.060206 | -0.10863 | 0.008311 | 0.124732 | 10294 | 0.99966 |
| twice_lag_irritability | -0.14139 | 0.071009 | -0.28254 | -0.14091 | -0.00241 | 11335 | 0.999707 |
| twice_lag_overall_mood | 0.138541 | 0.077537 | -0.01334 | 0.139106 | 0.293672 | 10516 | 0.999935 |
| is_weekend | 0.041827 | 0.070135 | -0.09386 | 0.041611 | 0.177111 | 13301 | 0.999717 |
| is_lag_weekend | -0.08866 | 0.072078 | -0.2314 | -0.08853 | 0.053009 | 12292 | 0.999609 |

**Table S12.** Fixed effect regression coefficients when targeting tiredness on sleep quality.

| | **Mean** | **SD** | **Q2.5** | **Median** | **Q97.5** | **N_eff** | **Rhat** |
|---|---|---|---|---|---|---|---|
| (Intercept) | -0.04157 | 0.477952 | -0.98979 | -0.04414 | 0.898226 | 14778 | 0.999747 |
| lag_tiredness | 0.006214 | 0.048785 | -0.08801 | 0.00562 | 0.101748 | 16572 | 0.999942 |
| lag_sleep_quality | -0.0343 | 0.051747 | -0.13621 | -0.0344 | 0.068515 | 15590 | 0.999631 |
| site | -0.01839 | 0.068865 | -0.15541 | -0.01883 | 0.115759 | 17344 | 0.999559 |
| gender | 0.041421 | 0.069432 | -0.09637 | 0.041914 | 0.176591 | 16229 | 0.999627 |
| age | 0.006293 | 0.020097 | -0.03324 | 0.006322 | 0.045576 | 13574 | 0.999888 |
| selection | -0.04332 | 0.072737 | -0.18245 | -0.04392 | 0.102677 | 16111 | 0.999969 |
| twice_lag_sadness | -0.15042 | 0.09635 | -0.34098 | -0.14936 | 0.040964 | 13372 | 0.999763 |
| twice_lag_anxiety | 0.026002 | 0.058756 | -0.08955 | 0.026571 | 0.141031 | 18372 | 0.999703 |
| twice_lag_inactivity | 0.081916 | 0.07699 | -0.06707 | 0.082257 | 0.231915 | 16507 | 0.999615 |
| twice_lag_tiredness | 0.006193 | 0.062004 | -0.11478 | 0.006408 | 0.125681 | 18009 | 0.999755 |
| twice_lag_irritability | -0.14647 | 0.068718 | -0.28187 | -0.14634 | -0.01221 | 17994 | 0.999551 |
| twice_lag_overall_mood | 0.131215 | 0.078454 | -0.02541 | 0.131002 | 0.284471 | 18308 | 0.999806 |
| is_weekend | 0.046381 | 0.07105 | -0.09222 | 0.046216 | 0.187276 | 16988 | 0.999807 |
| is_lag_weekend | -0.08984 | 0.073007 | -0.23393 | -0.08963 | 0.053731 | 18083 | 0.999983 |

**Supplement 6. Lagged Effects of Sleep on Mood After Subtracting Prior Nights Sleep**

**Table S13.** Fixed effect regression coefficients when targeting the total effect of lagged sleep quality on anxiety.

| | Mean | SD | Q2.5 | Median | Q97.5 | N_eff | Rhat |
|---|---|---|---|---|---|---|---|
| (Intercept) | -0.5422 | 0.570095 | -1.68568 | -0.53885 | 0.556534 | 6419 | 0.999973 |
| lag_sleep_quality | -0.13432 | 0.062949 | -0.26026 | -0.13362 | -0.01407 | 7857 | 0.99982 |
| twice_lag_sadness | 0.114209 | 0.107024 | -0.09699 | 0.113213 | 0.32505 | 8241 | 1.0002 |
| twice_lag_anxiety | 0.076907 | 0.064969 | -0.04925 | 0.076539 | 0.205807 | 9791 | 1.000285 |
| twice_lag_inactivity | -0.19595 | 0.083921 | -0.36275 | -0.19646 | -0.03003 | 8967 | 0.999801 |
| twice_lag_tiredness | 0.126492 | 0.066663 | -0.00185 | 0.126912 | 0.257931 | 10670 | 0.999642 |
| twice_lag_irritability | 0.191894 | 0.07612 | 0.043746 | 0.192019 | 0.344095 | 11391 | 1.000102 |
| twice_lag_overall_mood | -0.17572 | 0.087319 | -0.34576 | -0.17479 | -0.0024 | 9354 | 0.999927 |
| twice_lag_sleep_quality | -0.10098 | 0.055689 | -0.21173 | -0.10074 | 0.008479 | 10060 | 0.999751 |
| site | 0.0106 | 0.078467 | -0.14157 | 0.010398 | 0.167457 | 9514 | 0.999829 |
| gender | 0.002949 | 0.083494 | -0.16203 | 0.002822 | 0.167801 | 8337 | 1.000207 |
| age | 0.028078 | 0.023756 | -0.01655 | 0.027628 | 0.076576 | 7095 | 0.999942 |
| selection | 0.035691 | 0.082696 | -0.12422 | 0.036078 | 0.197386 | 9183 | 0.999913 |
| is_weekend | -0.04614 | 0.074726 | -0.19151 | -0.04753 | 0.100419 | 11179 | 0.99973 |
| is_lag_weekend | -0.02376 | 0.078786 | -0.17837 | -0.02545 | 0.134901 | 12428 | 0.999597 |

**Table S14.** Fixed effect regression coefficients when targeting the total effect of lagged sleep quality on irritability.

| | Mean | SD | Q2.5 | Median | Q97.5 | N_eff | Rhat |
|---|---|---|---|---|---|---|---|
| (Intercept) | 0.663788 | 0.504229 | -0.3336 | 0.666315 | 1.64825 | 5909 | 1.000607 |
| lag_sleep_quality | -0.12225 | 0.054999 | -0.2313 | -0.12197 | -0.01562 | 7415 | 1.000965 |
| twice_lag_sadness | 0.300671 | 0.096363 | 0.114841 | 0.300084 | 0.493396 | 6822 | 0.999834 |
| twice_lag_anxiety | -0.05206 | 0.059554 | -0.16801 | -0.05083 | 0.062686 | 8211 | 1.000004 |
| twice_lag_inactivity | -0.14137 | 0.078462 | -0.29553 | -0.14226 | 0.0133 | 7595 | 0.999916 |
| twice_lag_tiredness | 0.075387 | 0.060703 | -0.04369 | 0.07424 | 0.193485 | 8703 | 0.999947 |
| twice_lag_irritability | 0.172318 | 0.068564 | 0.03763 | 0.171466 | 0.307273 | 9388 | 0.999839 |
| twice_lag_overall_mood | -0.2094 | 0.079179 | -0.36277 | -0.20962 | -0.05244 | 8490 | 0.999641 |
| twice_lag_sleep_quality | -0.07268 | 0.051001 | -0.17189 | -0.07213 | 0.02896 | 10419 | 0.999936 |
| site | -0.0023 | 0.072009 | -0.14466 | -0.00165 | 0.13893 | 9310 | 0.999808 |
| gender | -0.07687 | 0.076868 | -0.22387 | -0.07819 | 0.082085 | 7030 | 1.00085 |
| age | -0.02678 | 0.020818 | -0.0681 | -0.02679 | 0.013788 | 6900 | 1.000001 |
| selection | -0.02317 | 0.077009 | -0.16912 | -0.0236 | 0.131299 | 7125 | 1.000667 |
| is_weekend | 0.004325 | 0.069112 | -0.13177 | 0.003529 | 0.140758 | 8901 | 0.999781 |
| is_lag_weekend | -0.06445 | 0.072785 | -0.20639 | -0.06392 | 0.077677 | 9697 | 1.000012 |

**Table S15.** Fixed effect regression coefficients when targeting the total effect of lagged sleep quality on overall mood.

| | Mean | SD | Q2.5 | Median | Q97.5 | N_eff | Rhat |
|---|---|---|---|---|---|---|---|
| (Intercept) | -0.09166 | 0.492234 | -1.07419 | -0.09256 | 0.872805 | 11676 | 0.999757 |
| lag_sleep_quality | -0.09353 | 0.052505 | -0.19525 | -0.09308 | 0.007019 | 11901 | 0.999563 |
| twice_lag_sadness | 0.28942 | 0.096128 | 0.099731 | 0.287685 | 0.478171 | 12004 | 0.999693 |
| twice_lag_anxiety | -0.16649 | 0.060552 | -0.28335 | -0.16723 | -0.04892 | 12270 | 0.999769 |
| twice_lag_inactivity | -0.03703 | 0.076919 | -0.18774 | -0.03546 | 0.111207 | 12262 | 0.99971 |
| twice_lag_tiredness | 0.009758 | 0.060122 | -0.10638 | 0.009341 | 0.130349 | 14457 | 1.000054 |
| twice_lag_irritability | 0.051113 | 0.068869 | -0.08306 | 0.05101 | 0.185194 | 12889 | 0.999629 |
| twice_lag_overall_mood | 0.01607 | 0.079342 | -0.1391 | 0.016245 | 0.171438 | 13344 | 0.999908 |
| twice_lag_sleep_quality | -0.06712 | 0.049847 | -0.16537 | -0.06657 | 0.032173 | 12929 | 0.999838 |
| site | -0.05113 | 0.066317 | -0.18009 | -0.05149 | 0.077056 | 11995 | 0.999654 |
| gender | -0.01281 | 0.071483 | -0.15145 | -0.01297 | 0.128082 | 13169 | 0.999647 |
| age | 0.006399 | 0.020502 | -0.03414 | 0.006467 | 0.047448 | 11832 | 0.999889 |
| selection | 0.058072 | 0.072962 | -0.08361 | 0.058133 | 0.200132 | 10966 | 0.999655 |
| is_weekend | 0.016534 | 0.068597 | -0.1166 | 0.016298 | 0.150732 | 13318 | 0.999836 |
| is_lag_weekend | 0.005737 | 0.071786 | -0.13176 | 0.006293 | 0.14538 | 13513 | 0.999754 |

**Table S16.** Fixed effect regression coefficients when targeting the total effect of lagged sleep quality on inactivity.

| | Mean | SD | Q2.5 | Median | Q97.5 | N_eff | Rhat |
|---|---|---|---|---|---|---|---|
| (Intercept) | 1.053594 | 0.541746 | -0.00424 | 1.060166 | 2.12069 | 11845 | 0.999774 |
| lag_sleep_quality | -0.10073 | 0.058565 | -0.21639 | -0.10002 | 0.013147 | 14212 | 0.999925 |
| twice_lag_sadness | 0.198527 | 0.108337 | -0.00862 | 0.197232 | 0.411957 | 10283 | 0.999953 |
| twice_lag_anxiety | -0.20282 | 0.06893 | -0.33565 | -0.2035 | -0.06568 | 16093 | 0.999763 |
| twice_lag_inactivity | 0.130622 | 0.087272 | -0.04267 | 0.130333 | 0.300505 | 12372 | 0.999627 |
| twice_lag_tiredness | 0.05494 | 0.068949 | -0.08019 | 0.054764 | 0.189577 | 14248 | 0.999812 |
| twice_lag_irritability | 0.028866 | 0.076854 | -0.12265 | 0.028505 | 0.182937 | 15104 | 0.999597 |
| twice_lag_overall_mood | -0.10659 | 0.091009 | -0.28568 | -0.10571 | 0.069637 | 13880 | 0.999641 |
| twice_lag_sleep_quality | 0.003279 | 0.056858 | -0.10865 | 0.002687 | 0.116497 | 15179 | 0.999794 |
| site | -0.08175 | 0.076205 | -0.22963 | -0.08212 | 0.068992 | 15464 | 0.999575 |
| gender | -0.11754 | 0.081212 | -0.2757 | -0.11835 | 0.043918 | 15248 | 0.999903 |
| age | -0.03789 | 0.022632 | -0.08147 | -0.03793 | 0.006967 | 12611 | 0.999669 |
| selection | -0.04609 | 0.083557 | -0.20655 | -0.04671 | 0.121453 | 14020 | 1.00014 |
| is_weekend | 0.021529 | 0.079818 | -0.13507 | 0.022416 | 0.179463 | 13623 | 0.999779 |
| is_lag_weekend | 0.02381 | 0.082602 | -0.14052 | 0.024928 | 0.184793 | 14949 | 0.999638 |

**Table S17.** Fixed effect regression coefficients when targeting the total effect of lagged sleep quality on sadness.

| | Mean | SD | Q2.5 | Median | Q97.5 | N_eff | Rhat |
|---|---|---|---|---|---|---|---|
| (Intercept) | 0.413846 | 0.538694 | -0.67086 | 0.41973 | 1.445583 | 5860 | 1.000458 |
| lag_sleep_quality | -0.13512 | 0.058305 | -0.25117 | -0.1341 | -0.02367 | 9072 | 0.999892 |
| twice_lag_sadness | 0.237075 | 0.101054 | 0.042084 | 0.236211 | 0.437607 | 8612 | 0.999833 |
| twice_lag_anxiety | -0.14725 | 0.06374 | -0.27185 | -0.1473 | -0.02375 | 12760 | 1.000192 |
| twice_lag_inactivity | 0.087225 | 0.082547 | -0.07696 | 0.087862 | 0.248422 | 7728 | 0.999847 |
| twice_lag_tiredness | 0.023753 | 0.066049 | -0.10665 | 0.023375 | 0.156466 | 8908 | 0.999988 |
| twice_lag_irritability | 0.040545 | 0.073235 | -0.10187 | 0.041038 | 0.182438 | 11002 | 0.999763 |
| twice_lag_overall_mood | -0.06397 | 0.08566 | -0.23358 | -0.06353 | 0.103663 | 9879 | 0.999767 |
| twice_lag_sleep_quality | -0.04595 | 0.053738 | -0.14909 | -0.04683 | 0.061082 | 12721 | 1.000336 |
| site | -0.08087 | 0.075582 | -0.23029 | -0.08081 | 0.070607 | 10245 | 0.999892 |
| gender | -0.07294 | 0.08132 | -0.2309 | -0.07342 | 0.087085 | 6334 | 0.999772 |
| age | -0.00786 | 0.022542 | -0.05143 | -0.00812 | 0.036935 | 6838 | 1.0002 |
| selection | -0.00859 | 0.081699 | -0.16403 | -0.01076 | 0.15839 | 8833 | 1.000144 |
| is_weekend | 0.036122 | 0.074477 | -0.10776 | 0.035374 | 0.181248 | 11259 | 0.999663 |
| is_lag_weekend | 0.005552 | 0.076524 | -0.14518 | 0.005061 | 0.153071 | 10674 | 0.999895 |

**Table S18.** Fixed effect regression coefficients when targeting the total effect of lagged sleep quality on tiredness.

| | Mean | SD | Q2.5 | Median | Q97.5 | N_eff | Rhat |
|---|---|---|---|---|---|---|---|
| (Intercept) | 1.099805 | 0.554147 | 0.005846 | 1.098095 | 2.194355 | 8702 | 0.999787 |
| lag_sleep_quality | -0.17056 | 0.057964 | -0.28587 | -0.17122 | -0.05698 | 10367 | 0.999879 |
| twice_lag_sadness | 0.073289 | 0.102775 | -0.12917 | 0.075022 | 0.27552 | 8052 | 1.000406 |
| twice_lag_anxiety | -0.2205 | 0.064612 | -0.34476 | -0.22021 | -0.09562 | 10943 | 0.99999 |
| twice_lag_inactivity | 0.006079 | 0.084214 | -0.15654 | 0.0064 | 0.171389 | 9485 | 1.000656 |
| twice_lag_tiredness | 0.235942 | 0.065659 | 0.10578 | 0.236768 | 0.363604 | 10357 | 1.0001 |
| twice_lag_irritability | 0.147184 | 0.075406 | -0.00043 | 0.14856 | 0.295508 | 11779 | 0.999833 |
| twice_lag_overall_mood | -0.16017 | 0.087226 | -0.33138 | -0.16097 | 0.010701 | 10476 | 0.999996 |
| twice_lag_sleep_quality | -0.04286 | 0.055207 | -0.15056 | -0.043 | 0.065798 | 13602 | 0.999724 |
| site | -0.06355 | 0.077118 | -0.2133 | -0.06456 | 0.088272 | 11274 | 0.999697 |
| gender | -0.05283 | 0.082208 | -0.21407 | -0.05338 | 0.111976 | 8762 | 0.999904 |
| age | -0.04392 | 0.023289 | -0.09004 | -0.04389 | 0.001835 | 10242 | 0.999764 |
| selection | -0.09369 | 0.083272 | -0.2567 | -0.09362 | 0.071478 | 8291 | 0.999578 |
| is_weekend | -0.12084 | 0.075142 | -0.26694 | -0.12155 | 0.025148 | 12748 | 0.999785 |
| is_lag_weekend | 0.078196 | 0.079143 | -0.07538 | 0.079164 | 0.235338 | 12571 | 0.999846 |

**Supplement 7. Interaction Effects**

We tested for an interaction effect between the selection criteria and the lagged mood variables on sleep quality. The estimated interaction effects were within one standard deviation of zero for all mood parameters as shown in Table S19 and S21. We also compared model performance statistics using 10-fold cross validation with the help of the loo package in R (Vehtari et al., 2024, 2017) shown in Table S20 and S22. Below shows the comparison between models as measured by expected log-predictive density (ELPD). In all cases, the model without the interaction term outperformed the model with the interaction term or the difference in ELPD was within one standard deviation. Therefore, we didn't have evidence to include an interaction term within our final analysis.

**Table S19.** Summary of the posterior distribution for the interaction term for lagged mood on sleep quality.

| Interaction | Mean | MCSE | SD | 2.5% | 50% | 97.5% | N_eff | Rhat |
|---|---|---|---|---|---|---|---|---|
| Anxiety | -0.089 | 0.001 | 0.100 | -0.218 | -0.089 | 0.037 | 8833 | 1.000 |
| Sadness | -0.030 | 0.001 | 0.115 | -0.176 | -0.029 | 0.118 | 9608 | 1.000 |
| Inactivity | -0.028 | 0.001 | 0.107 | -0.162 | -0.029 | 0.107 | 8132 | 1.001 |
| Tiredness | -0.024 | 0.001 | 0.098 | -0.149 | -0.024 | 0.103 | 11132 | 1.001 |
| Irritability | -0.118 | 0.001 | 0.115 | -0.265 | -0.118 | 0.028 | 10541 | 1.000 |
| Overall Mood | 0.050 | 0.001 | 0.112 | -0.091 | 0.050 | 0.194 | 8575 | 1.000 |

**Table S20.** Comparison between models with (Model 2) and without (Model 1) an interaction term between lagged mood and selection for the high-risk cohort on sleep quality. We show the difference in ELPD, ELPD per model, and the effective number of parameters (Param), along with their standard error.

| | Model | ΔELPD | SE ΔELPD | ELPD | SE ELPD | Param | SE Param |
|---|---|---|---|---|---|---|---|
| Anxiety | 2 | 0.00 | 0.00 | -456.46 | 26.55 | 25.64 | 4.98 |
| | 1 | -2.42 | 2.8 | -458.88 | 26.83 | 27.95 | 4.94 |
| Sadness | 1 | 0.00 | 0.00 | -459.42 | 25.58 | 28.86 | 4.82 |
| | 2 | -2.08 | 2.61 | -461.5 | 25.24 | 32.28 | 4.70 |
| Inactivity | 1 | 0.00 | 0.00 | -463.1 | 25.95 | 36.1 | 6.82 |
| | 2 | -0.67 | 3.41 | -463.77 | 24.77 | 37.32 | 5.79 |
| Tiredness | 2 | 0.00 | 0.00 | -460.43 | 24.44 | 26.6 | 3.8 |
| | 1 | -2.09 | 2.56 | -462.52 | 24.88 | 28.33 | 3.92 |
| Irritability | 1 | 0.00 | 0.00 | -457.3 | 25.72 | 26.25 | 3.87 |
| | 2 | -1.68 | 2.89 | -458.98 | 25.75 | 28.05 | 3.93 |
| Overall Mood | 1 | 0.00 | 0.00 | -458.42 | 26.07 | 29.88 | 5.22 |
| | 2 | -11.67 | 2.81 | -470.09 | 26.94 | 42.28 | 6.18 |

**Table S21.** Summary of the posterior distribution for the interaction term for sleep quality on mood.

| Interaction | Mean | MCSE | SD | 2.5% | 50% | 97.5% | N_eff | Rhat |
|---|---|---|---|---|---|---|---|---|
| Anxiety | 0.050 | 0.001 | 0.118 | -0.101 | 0.051 | 0.198 | 8468 | 1.000 |
| Sadness | 0.064 | 0.001 | 0.114 | -0.082 | 0.063 | 0.209 | 13532 | 1.000 |
| Inactivity | 0.056 | 0.001 | 0.112 | -0.085 | 0.055 | 0.198 | 10943 | 1.000 |
| Tiredness | 0.216 | 0.002 | 0.128 | 0.053 | 0.220 | 0.376 | 7063 | 1.000 |
| Irritability | 0.152 | 0.001 | 0.117 | 0.005 | 0.155 | 0.297 | 9610 | 1.000 |
| Overall Mood | 0.126 | 0.001 | 0.099 | 0 | 0.126 | 0.253 | 12528 | 1.000 |

**Table S22.** Comparison between models with (Model 2) and without (Model 1) an interaction term between sleep quality and selection for the high-risk cohort on mood variables. We show the difference in ELPD, ELPD per model, and the effective number of parameters (Param), along with their standard error.

| | Model | ΔELPD | SE ΔELPD | ELPD | SE ELPD | Param | SE Param |
|---|---|---|---|---|---|---|---|
| Anxiety | 1 | 0.00 | 0.00 | -475.10 | 21.47 | 26.24 | 3.03 |
| | 2 | -2.32 | 2.42 | -477.43 | 21.54 | 28.80 | 3.39 |
| Sadness | 1 | 0.00 | 0.00 | -448.71 | 23.50 | 24.99 | 4.40 |
| | 2 | -8.31 | 2.94 | -457.02 | 24.12 | 34.17 | 5.55 |
| Inactivity | 2 | 0.00 | 0.00 | -480.84 | 20.67 | 26.33 | 3.43 |
| | 1 | -0.69 | 2.79 | -481.52 | 20.50 | 26.86 | 3.25 |
| Tiredness | 2 | 0.00 | 0.00 | -489.37 | 20.86 | 36.22 | 5.12 |
| | 1 | -0.42 | 3.88 | -489.79 | 21.35 | 42.14 | 6.83 |
| Irritability | 1 | 0.00 | 0.00 | -435.19 | 22.27 | 37.36 | 3.93 |
| | 2 | -6.51 | 3.27 | -441.70 | 23.31 | 40.56 | 4.54 |
| Overall Mood | 2 | 0.00 | 0.00 | -420.48 | 21.62 | 27.98 | 3.27 |
| | 1 | -2.01 | 3.12 | -422.49 | 22.23 | 28.96 | 4.11 |

**Supplement 8. References**


Hernán, M.A., Robins, J.M., 2020. Causal Inference: What If. Chapman & Hall/CRC, Boca Raton.

Loh, W.W., Ren, D., 2023. A Tutorial on Causal Inference in Longitudinal Data With Time-Varying Confounding Using G-Estimation. Adv Methods Pract Psychol Sci 6. https://doi.org/10.1177/25152459231174029

Pearl, J., Glymour, M., Jewell, N.P., 2016. Causal Inference in Statistics: A Primer. John Wiley & Sons, Ltd.

VanderWeele, T.J., 2019. Principles of confounder selection. Eur J Epidemiol 34, 211–219. https://doi.org/10.1007/s10654-019-00494-6

Vehtari, A., Gabry, J., Magnusson, M., Yao, Y., Bürkner, P.-C., Paananen, T., Gelman, A., 2024. loo: Efficient leave-one-out cross-validation and WAIC for Bayesian models.

Vehtari, A., Gelman, A., Gabry, J., 2017. Practical Bayesian model evaluation using leave-one-out cross-validation and WAIC. Stat Comput 27, 1413–1432. https://doi.org/10.1007/s11222-016-9696-4
